# CSF Proteomics and Machine Learning Reveal Distinct Stages Across the Alzheimer’s Disease Continuum

**DOI:** 10.1101/2025.11.05.25339138

**Authors:** Saima Rathore, Eric B. Dammer, Anantharaman Shantaraman, Fang Wu, Duc M. Duong, Edward J. Fox, Erik C.B. Johnson, James J Lah, Alzheimer’s Disease Neuroimaging Initiative, Nicholas T. Seyfried, Allan I. Levey

## Abstract

Alzheimer’s disease (AD) is a neurodegenerative disorder characterized by heterogeneous pathophysiological changes that begin years before symptoms emerge. Existing biomarkers like Aβ and pTau capture only fragments of this complexity, limiting diagnosis and therapeutic development. Leveraging high-resolution cerebrospinal fluid (CSF) proteomics, quantifying 2,492 proteins using tandem-mass-tag mass spectrometry (TMT-MS), in 1,104 ADNI participants, we identified pathways reflecting AD pathogenesis and stage-specific molecular events *in-vivo*. In biomarker-positive MCI (due-to-AD) and AD Dementia, beyond well-established metabolic and mitochondrial dysfunction, we observed upregulated neuropeptide signaling, G-protein-coupled receptors activity, and synaptic remodeling, highlighting underrecognized synaptic and signaling alterations. Asymptomatic AD showed significant alterations in mitoschondrial metabolism, RNA processing, and extracellular matrix pathways. Across the continuum from asymptomatic AD to MCI (due-to-AD) and AD Dementia, 92 proteins were differentially abundant, revealing a stage-specific progression, with early disruptions in neurodevelopmental and extracellular vesicle-related pathways in asymptomatic and MCI (due-to-AD) participants, transitioning to impairments in intracellular signaling, synaptic architecture, and cytoskeletal integrity in AD Dementia. This progressive dysregulation supports a continuum model where early compensatory mechanisms gradually give way to widespread neuronal degeneration. Using machine learning, we derived CSF proteomic panels capable of accurately distinguishing disease stages (asymptomatic AD vs. MCI (due-to-AD): AUC=0.92; MCI (due-to-AD) vs. AD Dementia: AUC=0.87). In parallel, we developed machine learning models to estimate pathological burden (Aβ-PET, tau-PET), which substantially outperformed conventional biomarkers. These findings uncover protein signatures that reflect underlying AD biology and provide a foundation for stage-specific biomarkers and therapeutic targeting, with important implications for patient stratification and personalized intervention strategies.

**One Sentence Summary:** Comprehensive CSF proteomics across 1,104 ADNI participants delineated molecular signatures of Alzheimer’s disease pathogenesis and progression, enabling robust machine learning models for diagnosis, staging, and estimation of pathology.

## INTRODUCTION

Alzheimer’s disease (AD) represents a complex, multi-system disorder in which molecular disruptions unfold years before clinical symptoms appear (1–3). The earliest events involve imbalance in amyloid-beta (Aβ) metabolism, followed by the spread of tau pathology through vulnerable brain networks, ultimately converging on synaptic dysfunction and neuronal loss (2,3). This progressive biology blurs traditional diagnostic boundaries, leading to the recognition of AD as a continuum rather than a single clinical state. To capture this biological staging, the National Institute on Aging and Alzheimer’s Association introduced the A/T/N framework (4), which classifies individuals according to amyloid (A), tau (T), and neurodegeneration (N) biomarkers rather than cognitive status alone.

Building on this biological framework, recent advances in biomarker technology have transformed the detection and staging of AD pathology. Cerebrospinal fluid (CSF) biomarkers, including Aβ42, total tau, and phosphorylated tau (p-tau), now serve as cornerstone indicators of amyloidosis and tauopathy (5). The microtubule-binding region of tau has further proven to be a highly specific marker for neurofibrillary tangles, correlating strongly with tau-PET signal (6). Plasma-based biomarkers such as Aβ42/Aβ40 and p-tau isoforms (217, 181, 231) have expanded diagnostic reach beyond the clinic, showing performance comparable to CSF assays (7,8). While these biomarkers have greatly enhanced diagnostic precision and improved clinical trial stratification, they capture only part of AD’s molecular heterogeneity. Neuropathological studies demonstrate that AD frequently coexists with vascular injury, TDP-43 inclusions, or alpha-synuclein pathology in more than 90% of cases (9). Furthermore, biomarker positivity does not always parallel cognitive impairment: some symptomatic individuals show normal Aβ or tau levels, whereas up to 40% of cognitively unimpaired older adults exhibit underlying AD pathology (10,11). These inconsistencies highlight the need for additional molecular markers that capture complementary aspects of disease biology, enabling more accurate staging, monitoring of progression, and discovery of novel therapeutic targets.

Given the continued need for diverse AD biomarkers, our group and others have increasingly applied large-scale “omics” strategies to elucidate disease biology. Among these, proteomics provides a direct snapshot of the active molecular environment, complementing genomic and transcriptomic data to reveal downstream biological changes associated with disease progression (12–18). Through advances in mass spectrometry (MS)-based workflows, we have comprehensively profiled brain and CSF proteomes across hundreds of individuals, uncovering pathways linked to neurodegeneration, synaptic dysfunction, and glial activation (19–21). More recently, our CSF proteomic analyses identified numerous differentially abundant proteins across diagnostic groups in both sporadic and autosomal-dominant AD (22), underscoring the utility of

### MS proteomic signatures distinguish AD pathology [MCI (Due-to-AD) and AD Dementia] from AT- controls

To identify MS proteins reflective of AD pathology, we compared AT- controls to participants with AD Dementia or MCI (due-to-AD), enabling the discovery of proteins differentially abundant in symptomatic individuals with confirmed AD pathology. This analysis revealed 271 proteins with increased and 236 proteins with decreased abundance (P_FDR_<0.05; **Fig. 2A; table S3**), mapping to various biological pathways (**Fig. 2B**).

**Fig. 1.**
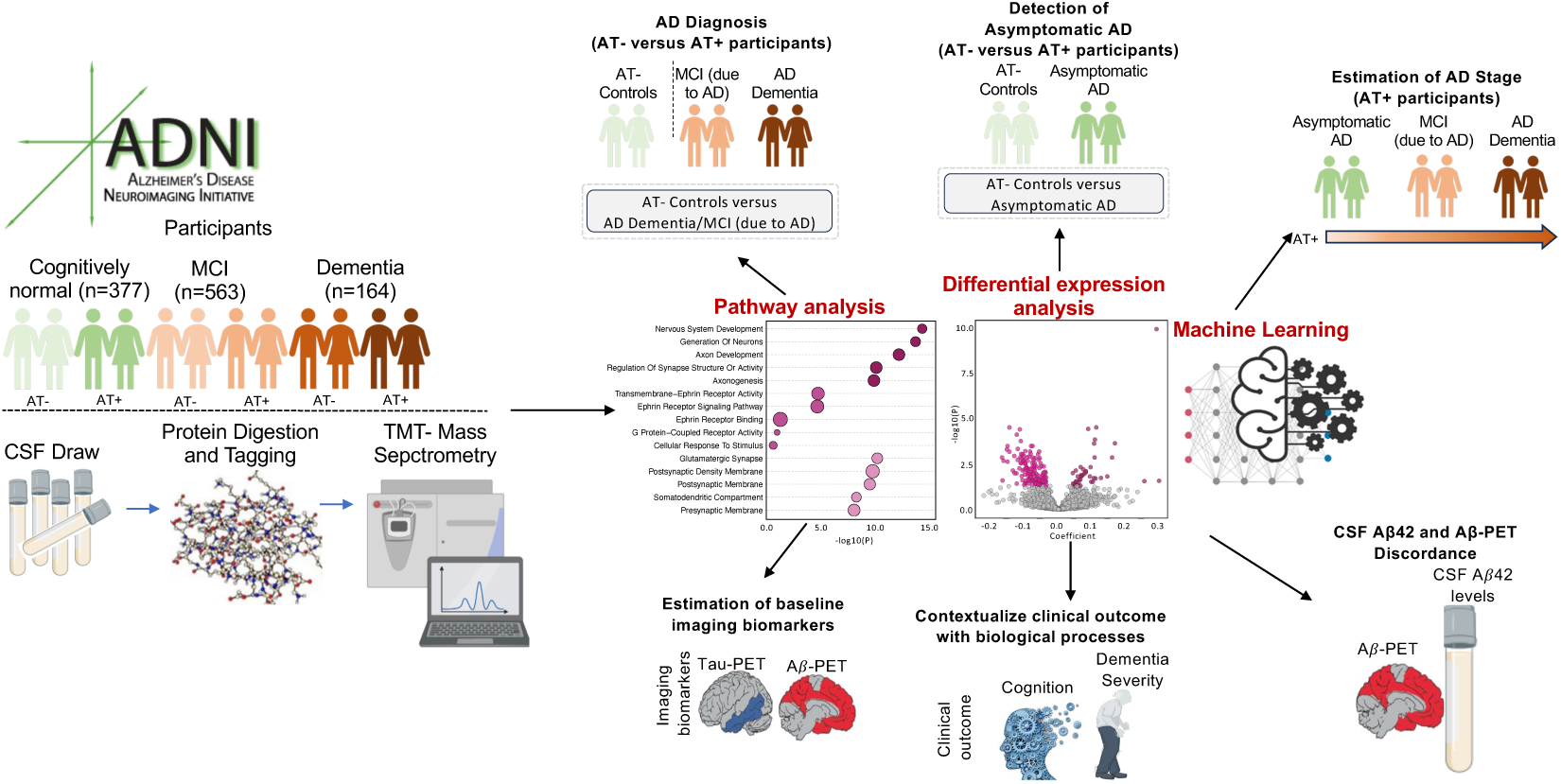
Study design overview. CSF was sampled from 377 controls [AT-=277, AT+=100], 563 MCI [AT-=259, AT+=304] and 164 Dementia [AT-=16, AT+=148] participants as defined by their clinical diagnosis and CSF Aβ42/pTau ratio (26). AT+ controls, MCI and Dementia participants will be referred as asymptomatic AD, MCI (due-to-AD), and AD Dementia. CSF proteomes were analyzed by tandem-mass-tag mass spectrometry (TMT-MS) to (i) identify differentially abundant proteins (DAPs) across disease groups, AD stages, and clinical or imaging measures, (ii) map them to Gene Ontology (GO) pathways, and (iii) train machine learning models for diagnosis, staging, and estimation of baseline pathological burden.

**Fig. 2.**
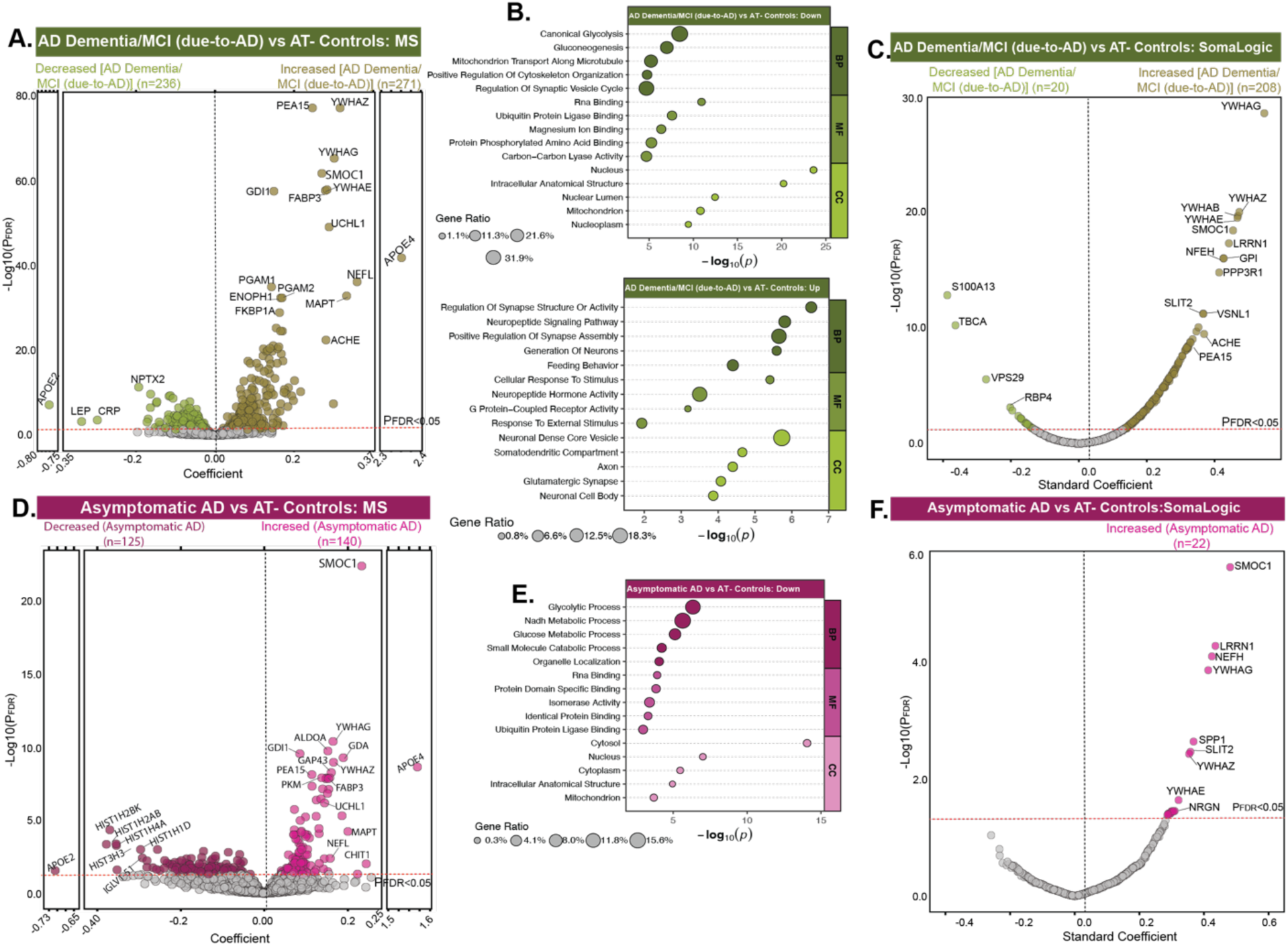
Differential abundance analysis across diagnostic groups. **(A)** Volcano plot depicting DAPs between AT- controls (n=277) and AD Dementia/MCI (due-to-AD) (n=452) using MS proteomics. **(B)** Summary terms from functional enrichment analyses using GO databases from the associations shown in A. **(C)** Corresponding analysis in SomaLogic cohort between AT- controls (n=108) and AD Dementia/MCI (due-to-AD) (n=292) for overlapping proteins (n=1,616). **(D)** Volcano plot depicting DAPs between AT- controls (n=277) and asymptomatic AD (n=100) using MS proteomics. **(E)** Summary terms from functional enrichment analyses using GO databases from the associations shown in D. **(F)** Corresponding analysis in SomaLogic cohort between AT- controls (n=108) and asymptomatic AD (n=33) for overlapping proteins (n=1,616). All P values were adjusted for FDR, and the red lines represent the threshold of P_FDR_<0.05, above which proteins were considered significant. Only the top proteins are labeled for legibility.

Twenty-four out of the 48 proteins from our earlier CSF targeted panel (23) were detected by MS analysis and 14 of those (ALDOA, APOE2, APOE4, DDAH1, GMFB, LDHB, NPTX2, NPTXR, PARK7, PEBP1, PPIA, SCG2, SMOC1, VGF) were highly significant (P_FDR_<0.05; **table S4**). Overall, significant changes included consistent increase of proteins involved in MAPK signaling and metabolism (PEA15, GPI), ubiquitin-proteasome system (UBE2V1, UCHL1), cytoskeletal organization (TAGLN3), neurotransmitter synthesis (PDXP), microtubule assembly (TBCA), metabolic processes (ENOPH1, PGAM), and intracellular trafficking (GABARAPL2). Several 14-3-3 proteins (YWHAZ, YWHAG, YWHAE) were also increased, consistent with their established roles in AD pathogenesis (21,27,28). Additionally, SMOC1, an extracellular matrix (ECM)-associated protein, was upregulated, supporting prior observations linking ECM remodeling to AD pathology in post-mortem brain tissue (12,13). In contrast, key synaptic proteins showed marked reductions, including VGF, a neurosecretory granin critical for synaptic vesicle transport, neurogenesis, and synaptic plasticity, whose decline reflects impaired neurotrophic support in AD (29). Similarly, NPTXR and NPTX2, neuronal pentraxins that regulate glutamatergic synaptic transmission and synapse formation, were significantly decreased, indicating progressive synaptic dysfunction and excitatory neurotransmission deficits characteristic of AD-related cognitive impairment (30,31). Beyond these established markers, several additional proteins showed significant changes, reflecting broader disruption of neuronal and synaptic integrity. Among these, PEA15 emerged as a particularly notable protein, previously shown to enhance astrocyte-mediated phagocytosis of Aβ and act as a protective factor against Aβ accumulation in AD models (32). Strikingly, PEA15 levels were found to change decades before clinical symptom onset, highlighting its potential utility as an early biomarker (33). Moreover, acetylcholinesterase (AChE), the enzyme responsible for acetylcholine breakdown and target of several FDA-approved cholinesterase inhibitors (34), was also increased. Other altered proteins included SHISA7, SORCS3, and CNR1 (synaptic regulation), DCC (axon guidance), JAG1 (Notch signaling), PCDHGC5 (cell adhesion), and PTPRN2 (metabolic and vesicle trafficking). The coordinated suppression of these pathways highlights an extended network of vulnerability in AD that spans beyond classical synaptic markers and may offer new opportunities for biomarker discovery and therapeutic targeting.

We replicated many of these differentially abundant MS proteins in 463 overlapping ADNI participants with CSF SomaLogic data, focusing on 1,616 proteins shared with MS (**Fig. 2C**). Of these, 416 (26%) and 566 (35%) proteins showed consistent negative and positive associations, respectively (P_FDR_<0.05; **table S5**), indicating strong cross-platform consistency for a substantial proportion of differentially abundant proteins (DAPs). In contrast, 634 proteins (39%) showed opposite associations, which may reflect platform-specific sensitivity or technical differences. Among 876 proteins uniquely quantified by MS, 60 were positively associated (P_FDR_<0.05), including biologically-relevant candidates such as APOE4, ENOPH1, NEDD8, CALM1, GLOD4, and CRYM, while 68 were negatively associated, including SHISA7, VGF, PTPRN2, MELTF, and CNR1. These findings highlight the value of integrating orthogonal proteomic technologies to validate AD biomarkers and reveal areas of potential measurement bias in indirect (e.g., aptamer-based) detection methods.

We next assessed the collective power of MS proteins in distinguishing AD Dementia and MCI (due-to-AD) from AT- controls. Using least absolute shrinkage and selection operator (LASSO)-based feature selection (35), we identified 17 key proteins (YWHAZ, SMOC1, PEA15, GDI1, UCHL1, APOE4, FABP3, NEFL, ACHE, NPY, MAPT, ENOPH1, PDYN, CRYM, CNR1, SELENBP1, CHI3L1) associated with the diagnostic groups. Random forest machine learning (36) model trained on the selected proteins of 80% of the data achieved median area under the receiver operating characteristic (AUC) curve of 0.97 (95% CI:0.95-1.00; **fig. S2; table S6**) across 100 permuted runs. Classification between clinically diagnosed controls and Dementia participants also showed high performance (median AUC=0.99; **fig. S3**). Notably, AChE consistently emerged as one of the top predictors, reinforcing its relevance to clinically diagnosed Dementia as well (**fig. S2C, S3I-J**). Overall, these analyses identified hundreds of altered proteins reflecting disease-relevant molecular changes, which could potentially serve as biomarkers of the disease.

### MS proteomic signatures of asymptomatic AD pathophysiology

Although cognitively normal, AT+ and AT- controls follow distinct biological trajectories. AT+ controls represent asymptomatic (preclinical) AD and carry higher risk for decline (4,37), while AT- controls show no biomarker evidence of AD. Identifying proteomic differences between these groups may offer insight into the earliest molecular changes in AD and highlight candidate pathways for secondary prevention trials.

Differential abundance analysis revealed 140 proteins with increased and 125 with decreased abundance (P_FDR_<0.05; **Fig. 2D; table S3**) in asymptomatic AD compared to AT- controls. To compare these findings with molecular changes previously characterized in autosomal dominant AD (ADAD), we examined 34 proteins identified as altered along the disease timeline in our DIAN study (22). This analysis recapitulated four of the five phases of molecular change observed in ADAD, detecting 30 of 34 proteins and confirming 19 with significant and directionally consistent alterations (**table S7**). Early alterations included extracellular matrix proteins (SMOC1, SPON1; stage-1), 14-3-3 proteins (YWHAZ, YWHAB, YWHAG) and glycolytic enzymes (PKM, LDHB, ALDOA, PGAM1; stage-2), followed by markers of axonal injury (NEFL; stage-3) and immune/metabolic stress (SPP1, CHI3L1; stage-4). In the final phase (stage-5), a second wave of glycolytic disruption (ENO2, PARK7) emerged, although synaptic and neurosecretory markers (SCG2, VGF, THY1, NPTX2, NPTXR), previously reported to decline after the estimated year of onset (EYO), had not yet changed. This was expected given that these changes were not evident before symptom onset. Notably, THY1 and ITGB2, which shift post-onset in ADAD, were already altered in asymptomatic sporadic AD, suggesting earlier engagement of synaptic and immune pathways. These results extend DIAN-based staging, revealing similar but earlier and broader molecular disruptions in sporadic AD, highlighting shared upstream biology across AD subtypes.

Beyond these established markers, we also identified several targets with marked changes in asymptomatic AD, including upregulation of GAP43, FABP3, CALM1, UCHL1, MAPT, CHIT1, and the decrease of histones (HIST1H2BK, HIST3H3, HIST1H2AB, HIST1H4A), and immunoglobulin proteins (IGHV1-2 and IGKV3-15) (P_FDR_<0.05). Overall, asymptomatic AD showed coordinated downregulation of pathways involved in cellular metabolism, RNA and protein regulation, and intracellular organization (**Fig. 2E**). These included glycolytic process, NADH and glucose metabolism, small-molecule catabolism, RNA binding, protein domain-specific binding, ubiquitin-protein ligase binding, and isomerase activity. Affected proteins localized mainly to cytosol, nucleus, mitochondria, and other intracellular compartments, suggesting suppression of essential homeostatic functions. This signature indicates that even prior to cognitive decline, asymptomatic individuals begin to show deficits in energy production, protein quality control, and intracellular trafficking, core processes maintaining neuronal integrity. These subtle, coordinated down-regulations likely reflect incipient neuronal stress or metabolic vulnerability associated with early AD pathophysiology and may serve as early molecular indicators of disease risk, preceding inflammatory or extracellular responses.

We replicated these differentially abundant MS proteins in 463 overlapping ADNI participants with CSF SomaLogic data, focusing on 1,616 proteins shared with MS (**Fig. 2F**). Sixty percent proteins showed consistent directionality across platforms (increased=638, decreased=341; **table S5**). Among 876 proteins uniquely quantified by MS, 25 were increased, including APOE4, GOT1, CALM1, NPEPPS, NEDD8, GLOD4, GOLIM4, and CRYM, while 56, primarily histones and immunoglobulins, were decreased (P_FDR_<0.05). LASSO-based feature selection and integrated random-forest model selected 8 key proteins (SMOC1, GDI1, GDA, APOE4, DNAJC3, CPVL, PCSKIN, PDYN) and showed median AUC of 0.91 (95% CI:0.84-0.98; **fig. S2B,D; table S8**) in distinguishing asymptomatic AD from AT- controls across 100 iterations of random subsampling in 80-20 train-test configuration.

### MS proteomic signatures of distinct AD stages in AT+ ADNI participants

Understanding how proteomic signatures evolve across clinical stages of AD individuals with confirmed AD pathology is critical for identifying molecular markers of progression and staging (38,39). We first assessed DAPs between asymptomatic AD and MCI (due-to-AD), and then between MCI (due-to-AD) and AD Dementia, to track protein changes along disease stages. A total of 142 proteins were differentially abundant (P_FDR_<0.05) between MCI (due-to-AD) and asymptomatic AD, of which 61.97% (n=88 proteins) were decreased in individuals with MCI (due-to-AD), including key proteins such as ADAMTS8, DCC, and ADGRB2, while 38.03% (n=54 proteins) were increased, with notable examples including PEA15, YWHAE, YWHAG, FABP3, ENOPH1, and AChE **(Fig. 3A).**

**Fig. 3.**
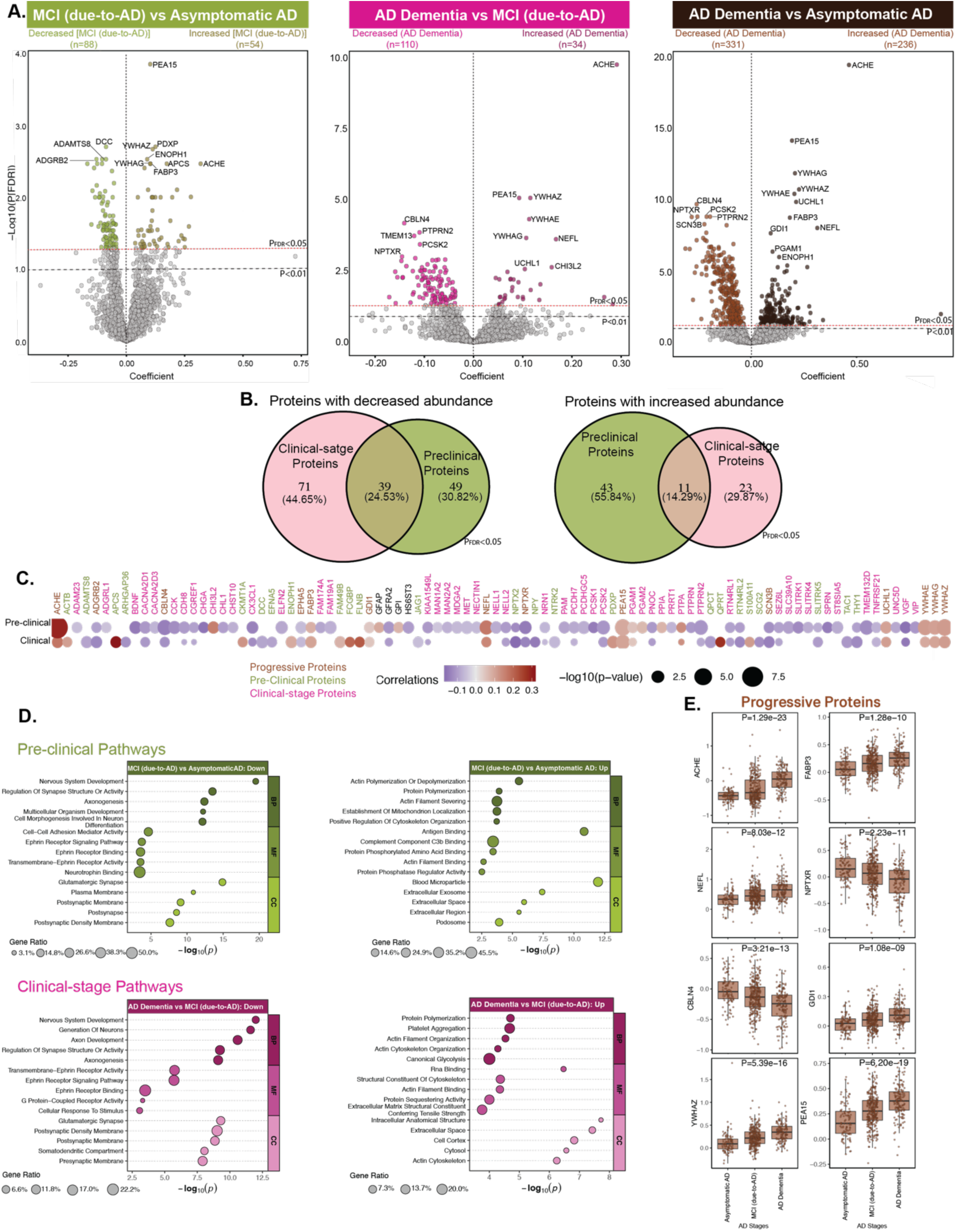
MS proteomic signatures across AD stages in AT+ ADNI participants. **(A)** TMT-MS data from individuals with confirmed AD pathology (asymptomatic AD: n=100; MCI (due-to-AD): n=304; AD Dementia: n=148) were analyzed. Volcano plots show DAPs for MCI (due-to-AD) versus Asymptomatic AD (Increased=54; Decreased=88), AD Dementia versus MCI (due-to-AD) (Increased=34; Decreased=110), and AD Dementia versus asymptomatic AD (Increased=236; Decreased=331). All P values were FDR adjusted (P_FDR_<0.05, red line). The x-axis shows the regression coefficient, the y-axis indicates -log10(P) value for all proteins. Only the top proteins are labeled for legibility. **(B)** Venn diagram summarizing overlap of DAPs across comparisons shown in **A** based on proteins significant at P_FDR_<0.05. **(C)** Heatmap of Pearson correlations between MS proteins and AD stages. Top proteins shown in **A** based on proteins significant at P_FDR_<0.01 were selected from each category, and their union is displayed in the heatmap. Proteins are labeled as their respective gene symbols, with colors indicating significance in pre-clinical or clinical AD stages. Dot color indicates correlation direction (red positive, purple negative), and size reflects -log10(P). **(D)** Summary terms from functional enrichment analyses using GO databases from the different categories of proteins. **(E)** Boxplots representing association of selected progressive proteins with AD stages.

Next, we studied which proteins changed with progression from MCI (due-to-AD) to AD Dementia stage. This comparison revealed 144 DAPs (P_FDR_<0.05), with 34 increased (23.61%) and 110 decreased proteins (76.39%) in individuals with AD Dementia (**Fig. 3A**). Overall, 52 proteins, including ACHE, ADGRB2, CBLN4, EPHA5, FABP3, GDI1, NEFL, NPTXR, PEA15, SCN3B, UCHL1, YWHAE, YWHAG, and YWHAZ were commonly altered across both comparisons, showing significant changes already in MCI (due-to-AD) participants and further deviation from control levels in the AD Dementia group (**Fig. 3B-C, E**). We refer to these as ‘progressive proteins,’ as they were differentially abundant across all AD stages and exhibited stage-dependent directional changes consistent with disease progression. DAPs specific to MCI (due-to-AD) versus asymptomatic AD and AD Dementia versus MCI (due-to-AD) are termed ‘pre-clinical’ and ‘clinical-stage’ DAPs, respectively. All summary statistics and protein panels are reported in **table S9-S10**.

In comparing protein pathway alterations between pre-clinical and clinical-stage AD (**Fig. 3D**), several pathways emerge as stage-specific. Down-regulated pathways exclusive to pre-clinical stage include those involved in neurodevelopmental processes (e.g., cell-cell adhesion, neurotrophin signaling, multicellular organism development), suggesting early disruptions in neuronal growth and connectivity. In contrast, clinical-stage down-regulation uniquely affects the somato-dendritic and presynaptic compartments, along with G-protein coupled receptor signaling, indicating deeper synaptic and signaling failure as disease progresses. Several pathways were consistently downregulated across both stages, including those involved in nervous system development, axonogenesis, synapse structure regulation, glutamatergic signaling, and ephrin receptor activity. This suggests that neuronal connectivity and synaptic integrity begin to deteriorate early in the disease and continue to decline with progression. Additionally, the emergence of presynaptic membrane, somato-dendritic compartment, and G-protein coupled receptor signaling exclusively in the clinical-stage down-regulated profile points to a worsening and broader spread of synaptic and signaling dysfunction over time, reflecting deepening neurodegeneration as AD advances.

In the preclinical stage, proteins increased in abundance were enriched in pathways related to actin remodeling, including filament severing and polymerization, mitochondrial localization, and cytoskeletal organization, alongside increased representation in extracellular compartments such as blood microparticles, exosomes, and podosomes (**Fig. 3D**). These findings suggest early, coordinated structural and vesicle-mediated responses to accumulating pathology, possibly reflecting immune activation, cytoskeletal adaptation, and intercellular communication. In contrast, clinical stage showed enrichment in platelet aggregation, canonical glycolysis, and broader cytoskeletal disorganization, with a shift in localization toward intracellular compartments such as the cytosol, mitochondria, and cell cortex. This transition from extracellular vesicle-related activity in early stages to intracellular structural breakdown in later stages underscores a spatial and functional progression of disease. Notably, protein polymerization and actin-related processes were shared across both comparisons, but their dysregulation intensified over time, evolving from limited remodeling in MCI to widespread disruption in AD. Together, these results support a continuum model of AD in which early compensatory and communicative processes give way to cell-intrinsic failure, marked by cytoskeletal collapse, metabolic stress, and impaired structural integrity.

### MS proteomics in combination with machine learning accurately detect AD stages

We performed 100 iterations of random subsampling with LASSO to identify key discriminative proteins. Proteins selected in >90% of iterations formed the pre-clinical (58 proteins; MCI (due-to-AD) vs. asymptomatic) and clinical-stage (16 proteins; AD dementia vs. MCI (due-to-AD)) discriminative panels. Notably, 43 proteins were exclusive to pre-clinical panel, 10 to the clinical panel (NEFL, MMP3, CBLN4, LECT2, ST8SIA5, SBSPON, GPNMB, VIP, RGN, PRND), and six (ACHE, PEA15, OPALIN, YWHAZ, MAPT, DNAJC5) overlapped, highlighting stage-specific signatures. Finally, the overall AD diagnostic panel (AD Dementia vs. asymptomatic AD) comprising 18 proteins was derived. Random forest model trained on these panels achieved median AUC of 0.92 (95% CI:0.85-0.99), 0.87 (95% CI:0.80-0.94), and 0.98 (95% CI:0.96-1.00) in distinguishing MCI (due-to-AD) from asymptomatic AD, AD Dementia from MCI (due-to-AD), and AD Dementia from asymptomatic AD (**Fig. 4A-C**).

**Fig. 4.**
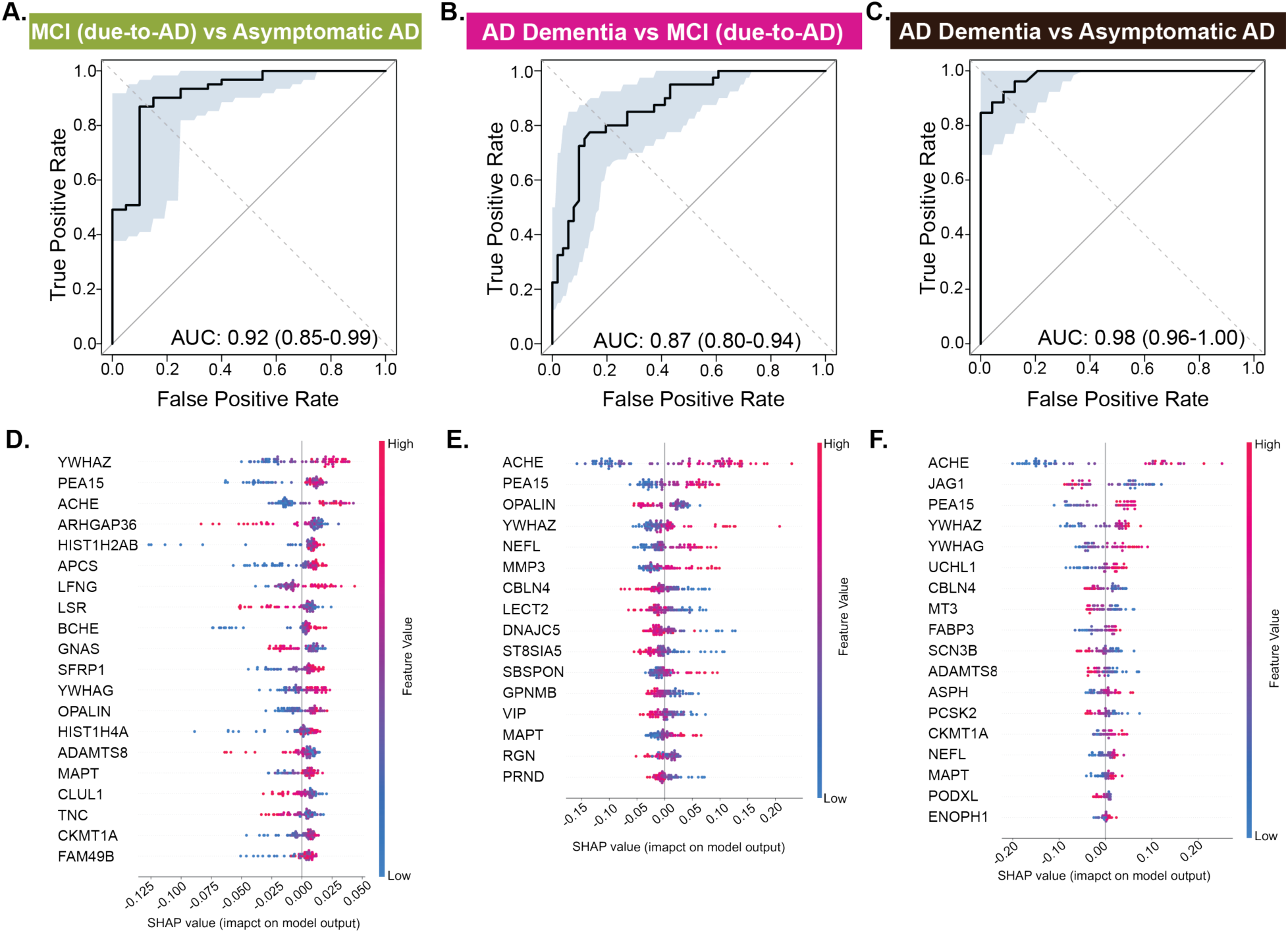
MS proteins associated with AD stages reveal predictive biomarkers for respective stages. **(A-C)** MS proteomics data were used to train classification models. LASSO based feature selection along with random forest classification-based model were trained. ROC curves from 100 permuted runs illustrate the median Area Under the Curve (AUC) based on an 80-20% train-test split for the following comparisons: (A) MCI (due-to-AD) versus asymptomatic AD (AUC=0.92), (B) AD Dementia versus MCI (due-to-AD) (AUC=0.87), (C) AD Dementia versus asymptomatic AD (AUC=0.98). Shaded area shows the worst and the best performance of the model across the 100 permuted runs. Higher AUC values indicate better classification performance, with values closer to 1.0 reflecting greater sensitivity and specificity. **(D-F)** SHAP based feature analysis is conducted on the models’ outcomes to determine contributions of different proteins in predictive modelling. Top SHAP feature contributions corresponding to the median AUC models shown in panels A-C are shown for: (D) MCI (due-to-AD) versus asymptomatic AD, (E) AD Dementia versus MCI (due-to-AD), and (F) AD Dementia versus asymptomatic AD. Each row represents a protein, and each dot reflects an individual’s data. The x-axis shows the SHAP value, indicating how strongly each protein influenced the model’s prediction. Dot color reflects protein abundance (red=high, blue=low). Proteins are ranked by overall impact, with ACHE, YWHAZ, and PEA15 among the most influential.

To interpret protein importance across AD stages, we applied SHapley Additive exPlanations (SHAP) analysis, which quantifies each protein’s contribution to model predictions (**Fig. 4D-F**). AChE had the strongest impact in predicting especially the later stages, with higher values linked to greater likelihood of AD Dementia. Other proteins involved include synaptic (YWHAZ, YWHAG), metabolism (PEA15), and protein folding (FABP3). Histone proteins such as HIST1H4A and HIST1H2AB tend to be important predictors in asymptomatic AD, consistent with prior studies reporting epigenetic dysregulation in preclinical phase of AD (40). Conversely, NEFL (41), key protein involved in axonal integrity pathways, and RGN (42), a regulator of calcium signaling and mitochondrial homeostasis, become increasingly important in predicting clinical stages of AD, reflecting progressive neuronal cytoskeletal disruption and axonal degeneration. Given that AChE was a top predictor of AD Dementia and MCI (due-to-AD), we evaluated whether its elevation was driven by AChE inhibitor therapy or inherent to disease status. We found that AChE levels were consistently higher in clinically diagnosed MCI and Dementia and further elevated among those on AChE inhibitors, suggesting both disease- and treatment-related effects (**fig. S4**). Overall, MS-based proteomics captures biologically distinct, disease-relevant changes beyond conventional CSF biomarkers, offering deeper insight into AD progression mechanisms.

### MS proteomics helps contextualize AD-linked cognitive deficit and dementia severity within biological processes

Cognitive deficit and dementia severity reflect the downstream consequences of complex and multifactorial biological processes that evolve over years. While these clinical outcomes provide direct insight into the impairment, they emerge only after substantial underlying neuropathology has developed. Core CSF biomarkers show limited correlation with clinical outcome, reducing their utility for staging and monitoring AD (43,44). Therefore, we need biomarkers to bridge the gap between molecular-level disruptions and observable clinical symptoms, revealing how shifts in brain biology ultimately manifest as clinical deterioration.

To uncover the biological drivers contributing to worsening clinical outcome, we examined associations between MS proteomic profiles and Clinical Dementia Rating scale Sum of Boxes (CDR-SB) and Alzheimer’s Disease Assessment Scale-Cognitive Subscale (ADAS-Cog). We identified 545 proteins (225 increased, 320 decreased) associated with the CDR-SB and 658 proteins (289 increased, 369 decreased) with ADAS-Cog11 (**Fig. 5A**, subset of proteins; **table S11**). Comparative analysis revealed both shared and distinct molecular correlates of these clinical measures (**Fig. 5A-C**). Individuals with greater impairment across both scores showed decreased abundance of proteins (NPTX2, VGF, NRN1, CHL1, EPHA5, and NELL2), which support synapse organization, axonogenesis, and neuroplasticity, highlighting synaptic dysfunction as a key driver of clinical deterioration. Similarly, proteins such as UCHL1, PCSK2, PEA15, and BDNF, which play roles in neurotransmitter release, synaptic vesicle cycling, and neurotrophic support, were also decreased in abundance, indicating a collapse in neuronal functional integrity. To contextualize these altered pathways within brain molecular architecture, we mapped the CDR-SB and ADAS-Cog–associated proteins onto previously defined brain protein co-expression modules (**Fig. 5D**) from our earlier study (12). This analysis revealed that proteins associated with worse clinical outcomes were significantly enriched in synaptic signaling modules as well as proteasome and protein homeostasis modules, suggesting that both synaptic dysfunction and impaired proteostasis contribute to cognitive and functional decline in AD.

**Fig. 5.**
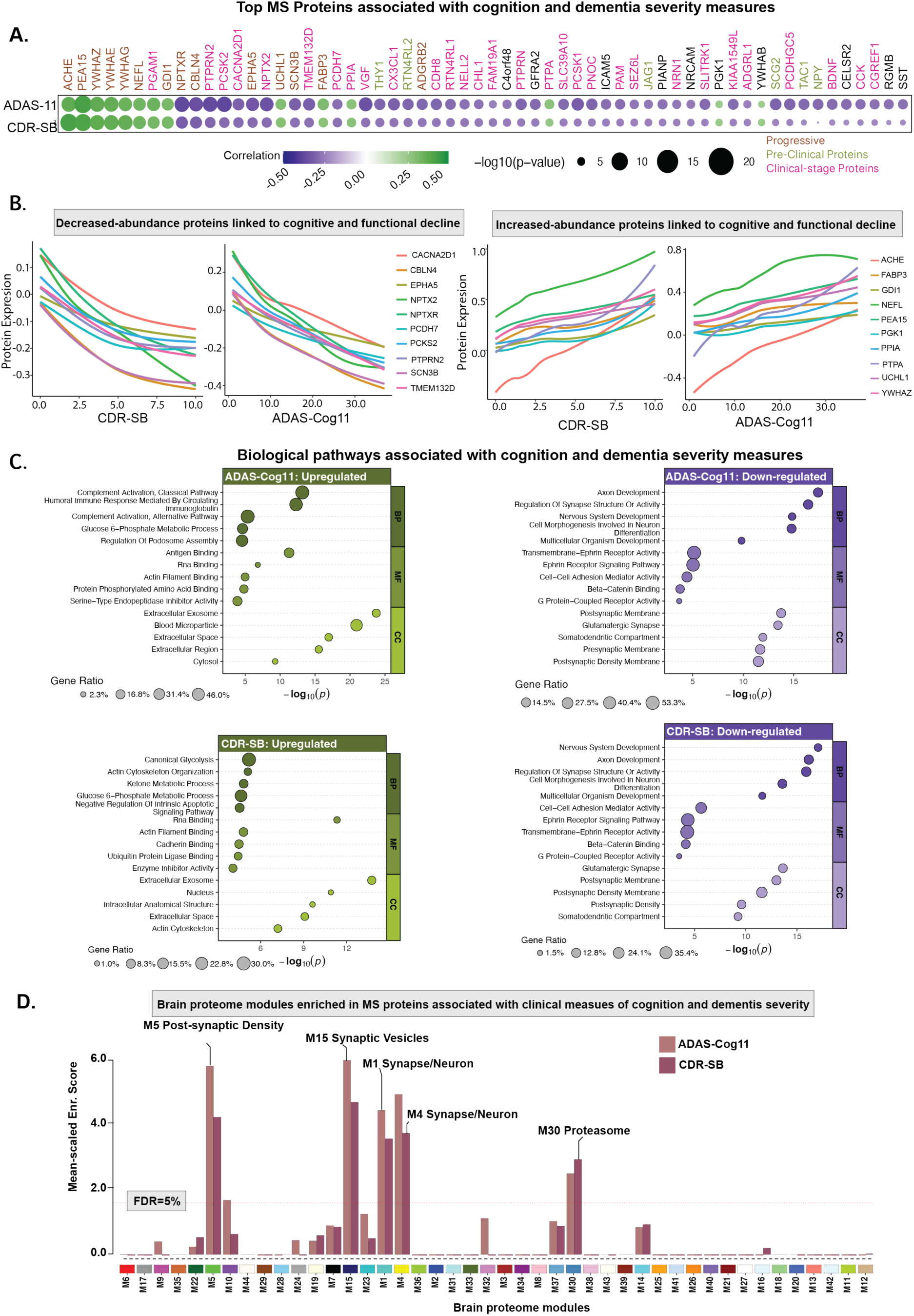
MS proteomics links AD-associated clinical outcomes with biological processes. **(A)** AT+ participants with ADAS-Cog11 (n = 551) and CDR-SB (n = 552) data were included. A heatmap displays Pearson correlations between MS protein abundance and clinical measures. Proteins significantly associated with either CDR-SB or ADAS-Cog11 (P_FDR_<0.01) were selected from each modality, and their union is shown. Each dot represents a correlation between protein levels and clinical measures, with gene symbols used for labeling. Protein names are color-coded based on whether they are significantly expressed in preclinical or clinical stages of AD. Dot color reflects the direction of correlation (green = positive, purple = negative), and dot size corresponds to the -log10(P). **(B)** Top 10 increased and decreased proteins selected from panel A based on the associations are shown. Smooth curve is fitted on the cross-sectional data showing association between clinical measures and specific protein abundance levels. **(C)** Summary of enriched biological processes derived from Gene Ontology analysis of the identified proteins. **(D)** MS proteins correlated with ADAS-Cog11 and CDR-SB scores (P<0.05) were mapped onto previously defined brain protein co-expression modules (12). The horizontal red dotted line marks the 5% FDR threshold from permutation testing, above which the enrichment was considered significant. Modules with significant enrichment or biologically relevant are labeled. Unlabeled but significant modules lacked defined annotations in the original network study.

Upregulated pathways diverged between cognitive decline and dementia severity (**Fig. 5C**). Cognitive decline (ADAS-Cog11) was primarily driven by immune-related processes, such as classical and alternative complement activation and humoral immune responses, with the involvement of several proteins including CX3CL1, PEA15, ICAM5, SERPINA3, and PPIA, while CDR-SB was enriched for metabolic and structural pathways, including glycolysis, ketone metabolism, cytoskeletal organization, and regulation of apoptosis. These distinct, protein-defined biological signatures, undetectable by conventional CSF markers and ADAS-11 and CDR-SB scores, demonstrate the added value of deep proteomic profiling in explaining the mechanisms underlying clinical outcome.

### MS proteomics helps contextualize neurodegeneration-linked imaging changes within biological processes

Neuroimaging measures such as FDG-PET, which assesses cerebral glucose metabolism, and structural MRI-derived hippocampal volume have been widely used to capture neurodegeneration (N in the A/T/N framework) (4). Although these imaging markers offer sensitive indicators of disease progression (45), they do not directly uncover the molecular mechanisms that drive them. To bridge this gap, we examined associations between MS proteomic profiles and FDG-PET standardized uptake value ratio (SUVR) and MRI-derived hippocampal volume (normalized by intracranial volume). This integrative approach allowed us to place imaging findings within a molecular context and link observable brain changes to biologically meaningful processes.

We found 988 proteins (465 increased, 523 decreased) associated with hippocampal volume, and 678 (361 increased, 317 decreased) with FDG-PET (P_FDR_<0.05; **Fig. 6A**, selected proteins are shown; **table S12**). The strongest associations included NPTX2 (R=0.39, P=7.22x10^-18^) and NPTXR with FDG-PET (R=0.39, P=8.88x10^-18^), and PCSK2 (R=0.43, P=3.96x10^-23^) and NPTX2 (R=0.42, P=2.53x10^-22^) with hippocampal volume. The regional analysis using cortical thickness revealed that associations were most prominent in the left superior temporal region (**Fig. 6B).** Overall, these proteomic associations revealed a consistent biological pattern linking neurodegeneration and metabolic decline with synaptic dysfunction and immune activation (**Fig. 6C**). These results suggest that as imaging markers reflect advancing neurodegeneration, a suppression of neuronal maintenance pathways occurs, evidenced by negative associations with proteins involved in axonogenesis (ROBO2, EPHA5), synapse organization (NPTX2, NPTXR, NRXN1), and cell morphogenesis (CHL1, DCC). Overlap analysis with brain proteome network modules revealed that these proteins aligned with modules associated with synapse organization and axonogenesis, mirroring the enrichment patterns observed in CSF (**Fig. 6D**). Moreover, analysis of biological pathways (**Fig. 6C**) further suggests that increased neurodegenerative burden on imaging is accompanied by heightened activity in immune and vascular pathways, with upregulated proteins enriched for complement activation (classical pathway), humoral immune response, blood coagulation, and wound healing. Notably, FDG-PET additionally showed enrichment for fibrinolysis, suggesting a more dynamic vascular response linked to metabolic stress, while hippocampal volume uniquely involved the alternative complement pathway, reflecting sustained innate immune activation. Many of these proteins (NPTX2, VGF, PCSK2, PEA15, CX3CL1) also overlapped with progressive or clinical-stage markers shown in **Fig. 3C**, indicating that these molecular alterations closely parallel clinical progression and likely emerge during symptomatic stages of disease.

**Fig. 6.**
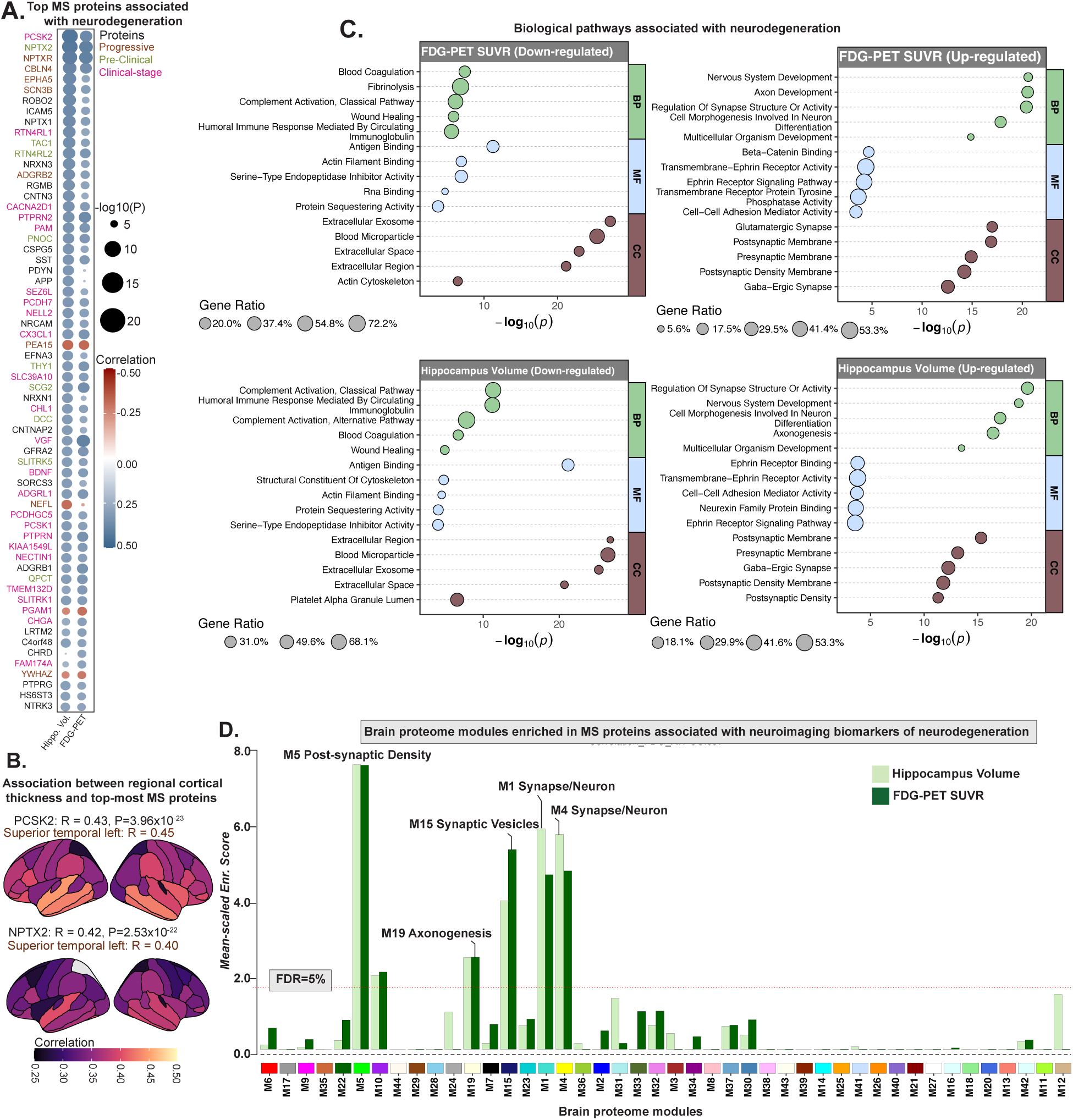
MS proteomics links AD-associated neuroimaging biomarkers of neurodegeneration with biological processes. **(A)** AT+ participants with available imaging biomarkers of neurodegeneration (FDG-PET SUVR [n=433] and hippocampal volume [n=478]) were included. A heatmap displays Pearson correlations between CSF protein levels and neuroimaging biomarkers. Proteins significantly associated with hippocampal volume or FDG-PET SUVR (top 40 proteins with P<0.01 and |Correlation (R)|>0.1 for each modality) were selected from each modality, and their union is shown. Each cell in the heatmap represents a correlation between protein levels and neuroimaging biomarkers, with gene symbols used for labeling. Color of each cell reflects the direction of correlation (red = positive, blue = negative). **(B)** Surface maps showing associations between top-ranked proteins from panel A and regional cortical thickness, highlighting widespread cortical atrophy in these participants. **(C)** Summary of enriched biological processes derived from Gene Ontology analysis of the identified protein categories. **(D)** MS proteins correlated with hippocampal volume and FDG-PET SUVR (P<0.05) were mapped onto previously defined brain protein co-expression modules (12). The horizontal red dotted line marks the 5% FDR threshold from permutation testing, above which the enrichment was considered significant. Modules with significant enrichment or biologically relevant are labeled. Unlabeled but significant modules lacked defined annotations in the original network study.

### MS proteomics accurately estimate Aβ and tau pathology, beyond conventional Aβ and tau biomarkers

Amyloid and tau PET imaging offers direct *in vivo* quantification of AD pathology and predicts future progression (46,47), but are costly, less accessible, and not routinely available in large or diverse clinical cohorts. In contrast, CSF proteomics captures a broad molecular snapshot of brain pathophysiology, including processes related to Aβ processing, tau phosphorylation, synaptic integrity, inflammation, and vascular injury. Therefore, modeling Aβ-PET and tau-PET using baseline CSF proteomics may enable cost-effective estimation of pathology in settings where PET is unavailable and reveal underlying biological pathways driving pathology.

Differential abundance analysis revealed 13 proteins (increased=8, decreased=5; P_FDR_<0.05) associated with [^18^F] AV45 (Aβ-PET) Centiloid value and 15 proteins (increased=13, decreased=2; P_FDR_<0.05) with [^18^F] AV1451 (tau-PET) neocortical SUVR (**Fig. 7A-B**; selected proteins are shown; **table S12)**. For tau-PET, proteins showing increased abundance included metabolic enzymes (PGAM1, PDXP, ENO1, ENO2, GPI, GDA), lysosomal proteins (UCHL1, PRCP), and cytosolic/exosomal markers (YWHAZ, YWHAE, YWHAG, FABP3, PDGFB) (**Fig. 7A-B**). These proteins were predominantly enriched in metabolic and lysosomal pathways, such as glycolysis, gluconeogenesis, catabolism, and carbohydrate/nucleoside biosynthesis, suggesting that increased tau burden is linked to metabolic reprogramming and vesicle-mediated processing. In contrast, decreased proteins included vascular and extracellular matrix regulators (PDGFB, EFNB1, TIMP3, SCUBE3, CHAD, VSIG4, SPOCK2). These proteins are associated with vascular and endothelial functions, including angiogenesis, blood vessel morphogenesis, and extracellular matrix signaling, indicating suppression of vascular remodeling in relation to tau accumulation (**fig. S5** shows biological pathways). Amyloid-PET showed similar trends (**Fig. 7A-B)**, where proteins with increased expression levels (PGAM1, PGAM2, ENO1, ENO2, GPI, MANBA, WFDC2, YWHAE, YWHAG, YWHAZ, PEA15, and SMOC1) reflected shifts in energy metabolism, glycoprotein catabolism, and cellular stress responses, with localization to intracellular compartments. Meanwhile, proteins with deceased expression levels (BDNF, CNTN5, CNTN3, SLIT3, PTPRN, PTPRN2, VGF, PNOC, DLK2, GFRA2, and EPHA10) were enriched in neuronal and synaptic pathways, including axon guidance, synapse organization, and neuropeptide signaling, and mapped to glutamatergic synapses and postsynaptic compartments (**fig. S5**). Together, these findings reveal a common molecular signature across tau and amyloid pathology: upregulation of metabolic and proteolytic systems, and downregulation of neuronal and vascular signaling, pointing to a disease-associated shift from structural and synaptic maintenance toward cellular stress and immune remodeling.

**Fig. 7.**
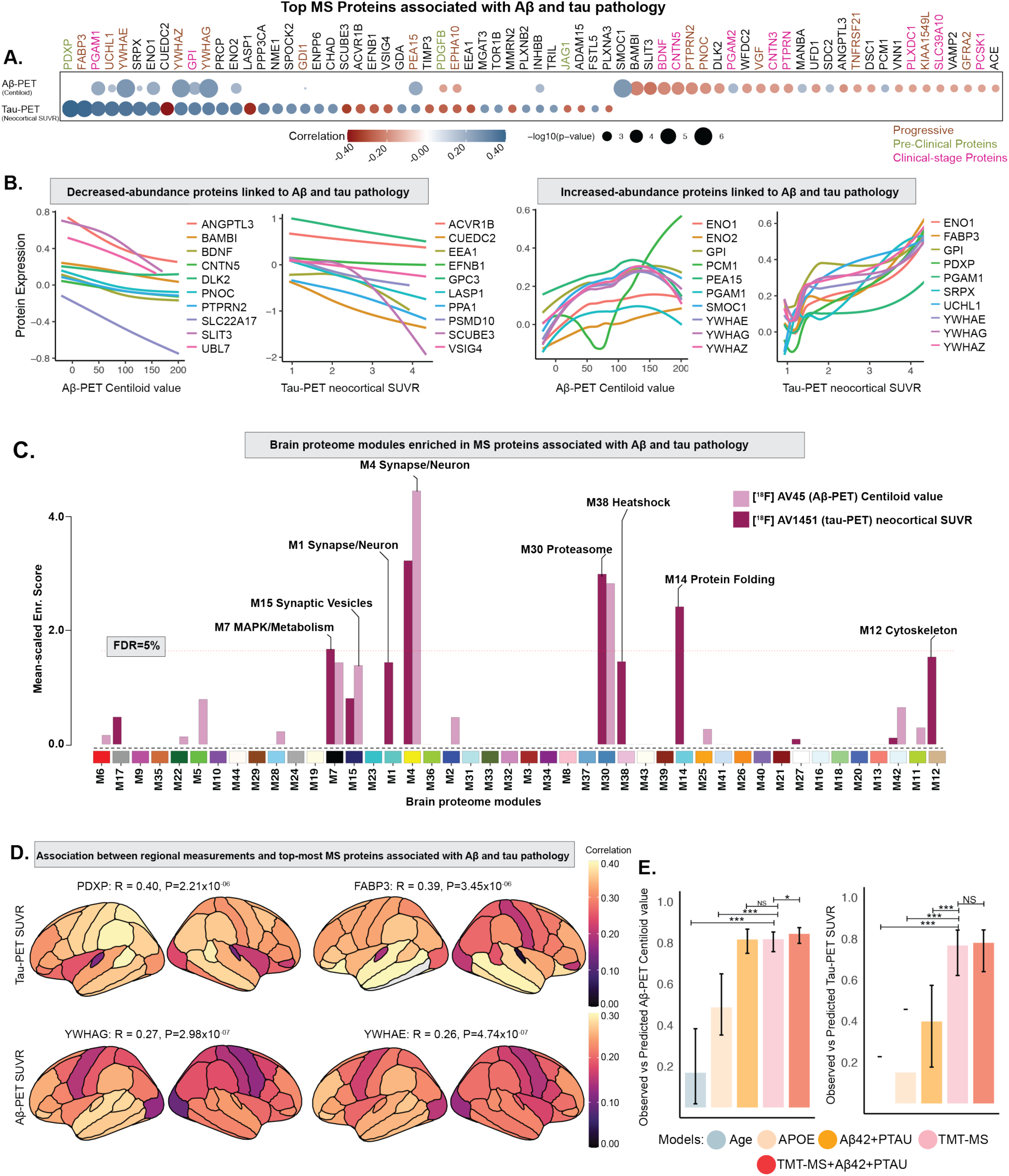
MS proteomics estimates baseline PET biomarkers, in AT+ participants beyond conventional Aβ and pTau biomarkers. **(A)** AT+ participants with available Aβ-PET and tau-PET images were included. A heatmap displays Pearson correlations between CSF protein levels and Aβ-PET based Centiloid value, and tau-PET neocortical SUVR. Proteins significantly associated with Aβ-PET Centiloid value and tau-PET SUVR (top 40 proteins with P<0.01 and |Correlation (R)|>0.1 for each modality) were selected from each modality, and their union is shown. Each cell in the heatmap represents a correlation between protein abundance levels and neuroimaging biomarkers, with gene symbols used for labeling. Color of each cell reflects the direction of correlation (red = positive, blue = negative). **(B)** Top 10 proteins with increased and decreased abundance levels, selected from panel A based on the associations, are shown. Smooth curve is fitted on the cross-sectional data showing association between PET biomarkers and specific protein levels. **(C)** MS proteins correlated with Aβ-PET based Centiloid value, and tau-PET SUVR (P<0.05) were mapped onto previously defined brain protein co-expression modules (12). The horizontal red dotted line marks the 5% FDR threshold from permutation testing, above which the enrichment was considered significant. Modules with significant enrichment or biologically relevant are labeled. Unlabeled but significant modules lacked defined annotations in the original network study. **(D)** Surface maps showing associations between top-ranked proteins from panel A and regional tau-PET SUVR and Aβ-PET SUVR, highlighting widespread pathology in these participants. **(E)** Random forest regression models were trained to estimate Aβ-PET Centiloid value and tau-PET SUVR. Bar plots show the Pearson correlation coefficients between observed and predicted values of PET biomarkers for models using the following predictors: 1) age alone, 2) APOE ε4 dose alone, 3) conventional AD CSF biomarkers including CSF Aβ42 and pTau as two features (“Aβ42+PTAU”), 4) CSF TMT-MS proteins alone (“TMT-MS”), and 5) CSF proteins and conventional biomarkers (“TMT-MS+Aβ42+PTAU”). (*p < 0.05, **p < 0.01, ***p < 0.001). NS = non-significant. Amyloid-PET (n=359) and tau-PET (n=133).

To evaluate how well the CSF proteome reflects AD pathology, we trained random forest regression models using MS protein levels following the same configurations described earlier and compared their performance with models based on age, APOE4 status, and conventional CSF biomarkers (Aβ42 and pTau181) (**Fig. 7E**). As expected, Aβ42 and pTau181 strongly predicted Aβ-PET Centiloid values (R=0.81, P=2.3×10^-32^) but showed weaker correlation with tau-PET SUVR (R=0.40, P=4.6×10^-04^). In contrast, MS-based proteomic models achieved slightly better performance for Aβ-PET (R=0.82, P=4.3×10^-33^) and significantly better performance for tau-PET (R=0.77, P=1.2×10^-15^), outperforming conventional biomarkers (P<0.001, permutation test). These gains likely reflect the proteome’s broader capture of synaptic, inflammatory, metabolic, and structural processes. Adding Aβ42 and pTau181 to the proteomic model yielded only minor improvements, indicating that proteomics encompasses most of the biologically relevant information in conventional biomarkers.

### MS proteomics reveals distinct biological pathways associated with CSF and PET biomarkers of AD

[^18^F] AV45 PET imaging has become a widely used *in vivo* marker of fibrillar amyloid pathology in AD (48), demonstrating strong concordance with CSF Aβ42 levels and reflecting a shared underlying disease process (27). However, a subset of individuals presents with discordant biomarker profiles, positive in one modality but negative in the other. These mismatches may reflect transitional stages of disease, alternative pathological mechanisms, or co-existing non-AD pathologies, underscoring the need for more granular biological profiling beyond standard Aβ biomarkers. Results of association testing of MS proteins with Aβ-PET (AV45) binding status (positive or negative) and CSF Aβ42 biomarker status (positive for CSF Aβ42<977 (44)) are shown in **Fig. 8A**. Aβ-PET binding was significantly associated with 174 proteins (32 with decreased and 142 with increased abundance levels; P_FDR_<0.01), while CSF Aβ42 was associated with a diverse array of 664 (394 with decreased and 270 with increased abundance levels; P_FDR_<0.01). Eighty-nine proteins were exclusively associated with Aβ-PET and 579 with CSF Aβ42, while 85 proteins showed an association with both (**Fig. 8A-B**).

**Fig. 8.**
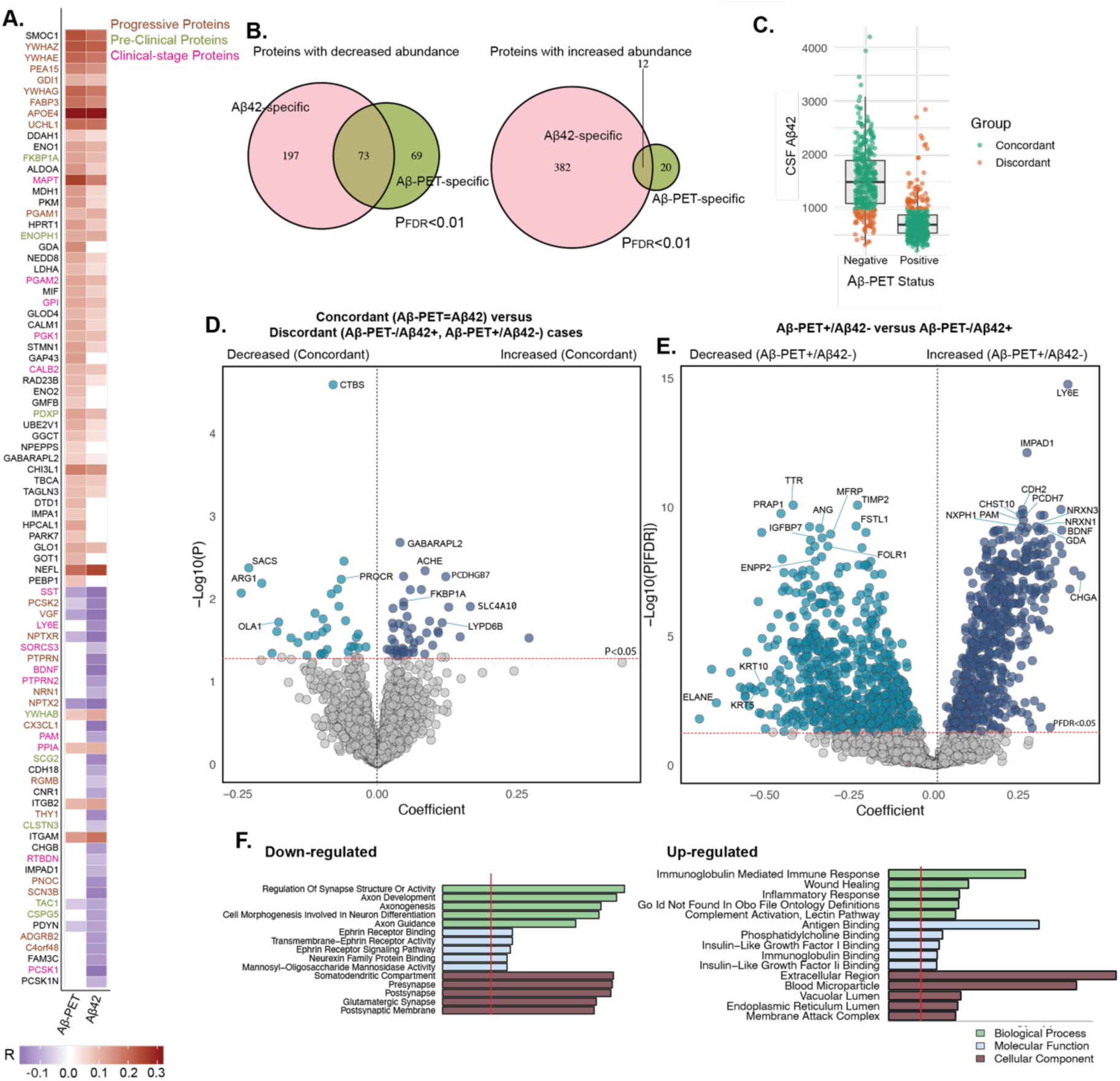
MS proteomics reveals insights into the discordance between Aβ pathology measured by Aβ-PET and CSF Aβ42 levels. **(A)** Samples with both Aβ-PET and CSF Aβ42 were included (n=745). A heatmap displays Pearson correlations between MS protein abundance levels and Aβ-PET or Aβ42 status. Heatmap shows top-most 50 associations with both Aβ-PET or Aβ42 (P<0.05). Each cell in the heatmap represents a protein-pathology correlation, with gene symbols used for labeling. Protein names are color-coded based on whether they are significantly expressed in preclinical or clinical stages of AD. Color of each cell reflects the direction of correlation (red = positive, purple = negative). **(B)** Venn diagram summarizing the overlap of proteins significantly associated (P<0.05) with Aβ-PET and Aβ42 across all participants. **(C)** Boxplot comparing participants with concordant Aβ pathology status (Aβ-PET-/Aβ42- or Aβ-PET+/Aβ42+; n=624) versus those with discordant status (Aβ-PET-/Aβ42+ or Aβ-PET+/Aβ42-; n=121). **(D)** Volcano plot shows differentially abundant proteins between concordant and discordant groups. The red line marks the significance threshold (P<0.05). The x-axis indicates regression coefficients, and the y-axis shows –log10(P). Only top-ranking proteins are labeled for readability. **(E)** Volcano plot shows differentially abundant proteins between Aβ-PET+/Aβ42- (n=61) and Aβ-PET-/Aβ42+ (n=60) participants. **(F)** Summary of enriched biological processes derived from Gene Ontology analysis of the protein categories identified in (E).

We looked deeper into the discordant cases by dividing them based on the positivity in each modality (Aβ42+/Aβ-PET-: n=60 and Aβ42-/Aβ-PET+: n=61; **Fig. 8C**). We observed upregulation of immune-related pathways in Aβ42-/Aβ-PET+ group, including immunoglobulin-mediated immune response, wound healing, inflammatory signaling, and complement activation, suggesting a potential immune-dominant molecular phenotype distinct from the classical Aβ-driven trajectory (**Fig. 8E-F**). This pattern is consistent with our recent proteomic studies that identified immune- and blood-brain barrier (BBB)-associated subtypes of AD (49). Notably, our findings also closely mirror the innate immune activation subtype reported in another recent CSF proteomics study (50), which showed broad activation of inflammatory and complement pathways.

These immune signatures may reflect not only distinct subtype but also a transitional stage, in which amyloid pathology is already detectable in the brain (Aβ-PET+) but not yet fully captured by CSF biomarkers. Together, these results support the existence of a biologically and clinically meaningful immune-dominant AD phenotype and reinforce the emerging molecular subtype framework identified through large-scale proteomic analyses.

## DISCUSSION

Despite decades of research, AD pathogenesis remains largely defined by Aβ and tau, yet these hallmarks capture only a fraction of the disease’s complexity. Given AD’s multifactorial nature, there is a pressing need for systems-level biomarkers that reflect its broader molecular and cellular underpinnings, enabling more precise and individualized interventions (34). Here, we applied a multi-dimensional framework integrating high-resolution MS-based CSF proteomics with clinical, cognitive, and neuroimaging data across the AD continuum. This approach uncovered AD-specific molecular pathways, stage-dependent protein alterations, and mechanisms underlying clinical and imaging outcomes. Machine learning applied to the proteomic data identified protein panels that accurately classified disease stages and estimated pathological burden.

Asymptomatic AD participants represent a critical stage for intervention, as cognition and brain integrity remain largely intact. In this early AD group, several MS proteins were already altered (**Fig. 2D-F**). Consistent with literature (22), SMOC1 increased with Aβ plaque load, even in cognitively-normal individuals, and was among the earliest proteins to change in ADAD and one of the strongest correlates of Aβ-PET in ADNI (23). In evaluating proteins previously altered in ADAD within our asymptomatic participants, we replicated four of the five defined molecular stages, confirming 19 of 33 key proteins across extracellular matrix, glycolytic, axonal, and immune pathways. Notably, proteins such as THY1 and ITGB2, which typically change years after symptom onset in ADAD, were already altered in asymptomatic sporadic AD, suggesting earlier molecular engagement in the disease process. The fact that nearly all significant changes found in ADAD (22) emerged in asymptomatic phase suggests that these early disruptions are shared across familial and sporadic AD subtypes. Another notable finding was the early and significant upregulation of PEA-15, a protein implicated in astrocyte-mediated Aβ clearance and stress responses, whose changes appearing decades before symptoms. Importantly, our analysis also identified several lesser-known or novel proteins with decreased abundance, including SHISA7 (inhibitory synaptic regulation), SORCS3 (intracellular trafficking of APP), CNR1 (neurotransmitter release), JAG1 (notch signaling) (51), and PCDHGC5 (cell adhesion), which, though not canonical AD biomarkers, are functionally linked to synaptic regulation, vesicle trafficking, neurotransmission, and signaling. Overall, these results reinforce and expand the temporal staging model (22) by capturing an earlier and broader molecular footprint of disease.

We also observed a marked rise in AChE levels, reinforcing that cholinergic dysfunction emerges well before cognitive symptoms, even though AChE has traditionally been studied only in symptomatic stages and treatment response. Notably, AChE expression was significantly higher in clinically-diagnosed dementia and MCI participants receiving cholinesterase inhibitors compared to untreated individuals (P=1.16×10⁻⁹; P<0.0001 after diagnosis adjustment). This pattern suggests treatment-induced upregulation of AChE, likely reflecting a compensatory response to enzymatic inhibition. Such feedback upregulation has been documented previously, where prolonged exposure to AChE inhibitors increased enzyme expression and activity as part of a homeostatic adaptation (52).

To better understand molecular cascade of AD, we examined proteomic changes exclusively in AT+ individuals, enabling direct comparison across asymptomatic, MCI (due-to-AD), and AD Dementia stages. We identified 52 progressive proteins whose abundance increased throughout the disease continuum (**Fig. 3E**). These proteins, several previously reported in AD (13,27), include ACHE, FABP3, NEFL, UCHL1, and YWHAZ, alongside others linked to neurodegeneration. Proteins like NPTXR, GDI1, SCN3B, YWHAE, and YWHAG are integral to synaptic function and intracellular signaling, while EPHA5, CBLN4, and ADGRB2 are implicated in cell adhesion, axon guidance, and neural connectivity. The 14-3-3 proteins are also increasingly reported as abundant in AD across studies (23). Notably, progressive proteins showed limited overlap with those altered between A^-^T^-^ and A^+^T^-^ participants and emerged later in the pathological cascade (A^+^T^-^ vs. A^+^T^+^), likely reflecting platform-specific differences in protein detection (53). Moreover, while that study characterized molecular events along the amyloid-tau axis, our analysis focused solely on AT+ participants stratified by clinical stage, enabling detection of proteomic changes linked to clinical progression rather than pathology accumulation alone.

Pathway analysis revealed stage-specific molecular shifts. Early-stage AD (MCI (due-to-AD) vs. asymptomatic AD) showed downregulation of neurodevelopmental and cell adhesion pathways, indicating early disruption of neuronal connectivity and plasticity. In contrast, late-stage transition (AD Dementia vs. MCI (due-to-AD)) involved deeper synaptic degeneration, with loss of presynaptic function and G-protein–coupled receptor signaling, reflecting advanced neuronal failure (54). Upregulated pathways also shifted from early extracellular and vesicle-mediated responses in MCI (e.g., actin remodeling, exosomes) to intracellular structural breakdown and metabolic stress in AD Dementia (e.g., glycolysis, cytoskeletal collapse). Consistent with prior studies (53), these glycolytic and metabolic disruptions intensified late in the disease, coinciding with widespread fibrillar pathology.

We developed two machine learning-based predictive models that yielded a 43-protein signature distinguishing preclinical stage and a 16-protein signature differentiating clinical stage, revealing distinct molecular landscapes of AD progression. Histone proteins (HIST1H4A and HIST1H2AB) emerged as key predictors in the preclinical stage, consistent with evidence linking histone methylation and acetylation changes to early AD pathophysiology (55). These epigenetic changes are thought to represent early molecular events preceding neurodegeneration, suggesting a role for chromatin remodeling in the initiation of AD-related gene expression programs. In contrast, NEFL, a neurofilament protein central to axonal integrity, was more predictive in clinical-stage AD, consistent with its role as a marker of axonal injury and disease progression (56). Its increased predictive value in advanced stages likely reflects progressive cytoskeletal breakdown and neuronal loss, hallmarks of symptomatic AD. Together, these results highlight a shift from early epigenetic dysregulation to later cytoskeletal disruption across the AD continuum.

A key finding of this study is the divergence in molecular signatures between CSF-/PET+ and CSF+/PET-individuals, revealing distinct underlying biology despite similar clinical classifications. Two complementary hypotheses may explain this pattern. First, the upregulation of immune pathways, including immunoglobulin response, wound healing, inflammatory signaling, and complement activation, suggests an immune-dominant AD subtype distinct from the classical Aβ-driven trajectory, consistent with reports of immune and BBB–related subtypes by recent large-scale proteomic studies (49,50). Second, these immune and synaptic signatures may reflect a transitional disease stage, a biologically active inflection point where fibrillar amyloid is already present in the brain (PET+), yet CSF Aβ42 remains within diagnostic thresholds. Proteomic profiling of CSF-/PET+ individuals revealed not only immune engagement and synaptic remodeling, but also a decline in neuroprotective and vascular-supporting proteins (e.g., TTR, FSTL1, ENPP2, ANG), suggesting increased vulnerability. Overall, these findings suggest that discordant biomarker profiles may reflect either distinct molecular subtypes or early, transitional stages of AD, with important implications for stratified diagnosis and intervention.

The proteomic insights from this study hold strong translational promise for both trials and clinical care. By capturing stage-specific molecular changes beyond Aβ and tau, our CSF proteomic panels provide a more comprehensive framework for characterizing AD. These data enable precise participant stratification, support early-stage trial enrollment, and offer protein signatures that predict clinical and imaging measures of disease burden, serving as potential surrogate endpoints. The findings also reveal novel, druggable pathways, such as synaptic signaling, neuropeptide activity, and ECM, expanding therapeutic targets. In clinical practice, these proteomic signatures could complement existing diagnostics, identify high-risk individuals, and enable personalized intervention and treatment monitoring, advancing precision medicine in AD.

While this study provides valuable insights into AD pathophysiology with strong translational relevance, several limitations remain. First, the ADNI cohort lacks population diversity, necessitating validation in more heterogeneous and racially diverse groups. Second, although blood-based tests would offer greater clinical utility, the translation of CSF protein signatures to plasma markers was not assessed. Nevertheless, replication of diagnostic proteins using SomaLogic platform supports their robustness across technologies. Overall, MS-based proteomics enhances A/T/N biomarkers by capturing broader AD-related mechanisms and improving diagnostic and staging accuracy.

Overall, this study presents a proteomic framework for identifying and validating systems-based CSF biomarkers for AD. Further optimization across cohorts and proteomic platforms may enhance risk stratification and therapeutic development, while longitudinal studies will clarify which marker combinations best track early risk and disease progression.

## MATERIALS AND METHODS

### Study design

The study was designed to identify CSF-based biological pathways reflective of AD pathogenesis, stage-specific molecular events, and clinical and neuroimaging measures of neurodegeneration and pathological burden, as well as to develop machine learning-based, robust biomarker panels indicative of diagnosis, disease stage, and underlying pathology. Data used in the preparation of this article were obtained from the Alzheimer’s Disease Neuroimaging Initiative (ADNI) database (adni.loni.usc.edu). The ADNI was launched in 2003 as a public-private partnership, led by Principal Investigator Michael W. Weiner, MD. The primary goal of ADNI has been to test whether serial magnetic resonance imaging (MRI), positron emission tomography (PET), other biological markers, and clinical and neuropsychological assessment can be combined to measure the progression of mild cognitive impairment (MCI) and early Alzheimer’s disease (AD). The ADNI study was approved by the Institutional Review Boards at each participating site (see full list here: http://adni.loni.usc.edu). All procedures were performed in accordance with relevant guidelines and regulations, and informed consent was obtained from all subjects prior to enrollment.

The current study was approved by the ADNI Data and Publications Committee (ADNI DPC). CSF collected at baseline visits were assayed from 1,104 participants recruited from ADNI. Samples were randomized and blinded for tandem-mass-tag mass spectrometry (TMT-MS) analyses. All participants had standardized diagnostic assessment that renders a clinical diagnosis of either control, MCI, or AD. Control participants had no subjective memory complaints, tested normally on Logical Memory II of Weschler Memory Scale (WMS-II), had an MMSE between 24-30, and a CDR of 0 with memory box score of 0. A subject is diagnosed as MCI if the study participant (i) reports concern due to impaired memory function; (ii) obtains a MMSE score between 24 and 30; (iii) a Clinical Dementia Rating Scale (CDR) score of 0.5; (iv) a score lower than expected (adjusted for years of education) on WMS-II; and (v) reports preserved function of daily living. AD participants exhibited subjective memory concerns and met NINCDS/ARDA criteria for probable AD. AD participants showed abnormal memory function on WMS-II, an MMSE of 20-26, and CDR of 0.5 or 0.1.

### Imaging acquisition and processing

This study includes Aβ ([^18^F] Florbetapir AV45) PET, tau ([^18^F] Flortaucipir AV1451) PET, and structural MRI (T1-weighted) scans obtained through the ADNI database. Preprocessed images from ADNI, with realigned frames, head position corrected through linear transformation, standardized voxel size, and smoothed to a uniform resolution of 6mm, were used in this study. Aβ-PET scans were analyzed in each participant’s native space, using their structural MRIs acquired closest to the Aβ-PET scan. The structural MRIs were segmented into cortical regions of interest and reference regions for each subject using FreeSurfer. The Aβ-PET data were then realigned, and the mean of all frames was used to co-register the Aβ-PET data with the corresponding structural MRI. For each subject, cortical SUVR images were generated by dividing voxel-wise florbetapir uptake by the average uptake from the whole cerebellum reference region.

For tau-PET data, a similar procedure was followed. The tau-PET images were realigned, and the mean of all frames was used to co-register the tau-PET data with the structural MRI closest in time. In each subject’s native MRI space, tau-PET SUVR images were generated by normalizing mean tau-PET uptake to an inferior cerebellar gray matter defined by spatially unbiased atlas template of the human cerebellum (57). The tau data used in this study is corrected for partial volume effects using the geometric transfer matrix approach (58).

To ensure consistency in our imaging assessments, we utilized the MRI, tau-PET, Aβ-PET, and FDG-PET scans obtained closest to the baseline CSF proteomics measurement. From these modalities, we extracted key biomarkers, including hippocampal volume from MRI, Centiloid value for Aβ-PET (59), tau-PET SUVR in the neocortex, and FDG-PET SUVR (averaged across the angular gyrus, temporal cortex, and posterior cingulate, regions known to exhibit early hypometabolism in AD). Hippocampal volume was normalized to intracranial volume to account for individual differences in brain size. Aβ-PET quantification followed the Centiloid Project framework, which standardizes Aβ burden measurement by defining a universal 0-100 scale, where 0 represents the mean SUVR of young controls and 100 corresponds to typical AD patients. Centiloid values were derived by linearly transforming tracer-specific SUVRs using the standardized PiB PET reference dataset (59). These imaging biomarkers were selected to provide a comprehensive representation of neurodegeneration, amyloid burden, and tau pathology, allowing us to investigate their associations with CSF proteomics profiles.

### CSF samples preparation and mass spectrometry

The overall workflow and data processing pipeline have been previously described in detail (60,61). Briefly, CSF proteins were digested and labeled using isobaric Tandem Mass Tag (5mg; TMTpro^TM^ 16-plex Label Reagent, Lot: # YA357799; 134C and 135N lot# YB370079, ThermoFisher Scientific) reagents following established protocols. Labeled peptides were pooled, desalted using mixed-mode cation exchange columns, fractionated under high-pH conditions, and analyzed by liquid chromatography-tandem mass spectrometry (LC-MS/MS) on an Orbitrap Astral spectrometer (ThermoFisher Scientific) equipped with a FAIMS Pro source. Data were processed using FragPipe (v22.0) with MSFragger (62) and Philosopher (63) for identification and quantification at a 1% FDR, searching against the UniProt human proteome supplemented with APOE and Aβ peptide sequences.

### Tunable approach for median polish of ratio (TAMPOR)

We used TAMPOR to remove technical batch variance in the MS data, as previously described (25). TAMPOR removes inter-batch variance while preserving variance caused by biological changes in the protein abundance values, normalizing to the median of selected samples.

### Regression of unwanted covariates

The protein abundance matrices are then subjected to non-parametric bootstrap regression by subtracting the trait of interest (batch of the sample) multiplied by the median estimated coefficient of fitting for each protein in the log2(abundance) matrix.

### CSF markers of Aβ42 and pTau181

CSF Aβ42 and pTau181 were measured using the Elecsys immunoassays (Roche Diagnostics). A pre-established cutoff of 39.20 on the CSF Aβ42/pTau181 ratio was used to define positivity on Aβ and tau pathologies (26). Participants below the cutoff were considered AT- (n=552; without a neurodegenerative disease diagnosis) and the ones above the threshold were considered as AT+ (n=552; with a neurodegenerative disease). Controls, MCI and Dementia in AT- group are referred as Controls, non-AD MCI, and non-AD Dementia, and the corresponding participants in AT+ group are referred as asymptomatic AD, MCI (due-to-AD), and AD Dementia.

### Differential protein expression analyses

Linear models were used to identify differentially abundant proteins (DAPs) across AD stages and between different diagnostic groups. The first set of comparisons focused on AD continuum: the asymptomatic AD versus MCI (due-to-AD) contrast captured early-stage changes, while the MCI (due-to-AD) versus AD Dementia contrast identified proteins associated with later stages of disease progression. The second set focused on comparing AT- controls with asymptomatic and symptomatic AD participants, including MCI (due-to-AD) and AD Dementia. Results are presented as volcano plots. To account for multiple hypothesis testing, we applied the Benjamini-Hochberg false discovery rate (FDR) method to adjust p-values.

### Enrichment analysis of brain network modules

In prior work, we identified 44 protein co-expression modules from deep proteomic profiling of over 1,000 dorsolateral prefrontal cortex samples, underscoring distinct proteomic architecture in AD. To link these brain-derived modules (M1-M44) (12) with CSF biomarkers of clinical and neuroimaging measurements, we tested CSF proteins associated with clinical and neuroimaging measures for enrichment within these modules. Gene-level p-values were assessed using 10,000 permutations in R (statmod::permp), and module-specific Z scores were calculated by comparing observed and permuted mean p-values. This approach mirrors the enrichment analysis applied to MAGMA- derived gene-level statistics for genome-wide association study, as described in previous work (12) (available at https://www.github.com/edammer/MAGMA.SPA).

### Machine learning modelling

To assess the predictive capability of MS proteins in distinguishing different stages of AD, we implemented a machine learning pipeline repeated over 100 iterations of random subsampling in an 80-20% split train-test configuration. First, feature selection was performed using the least absolute shrinkage and selection operator (LASSO) (35). Proteins selected in ≥90% iterations of random subsampling were retained as robust, stage-specific discriminative panels. Next, classification models were developed using the random forest (36) algorithm. Feature selection and hyperparameter tuning were conducted within the 80% training data using nested five-fold cross-validation. Model performance was evaluated using the area under the receiver operating characteristic curve (AUC). Machine learning analyses were conducted in Python (version 3.9), using libraries such as scikit-learn for model development and evaluation. Optimal thresholds along the ROC were defined using the Youden index (64). ROC curve analyses were performed in R using the pROC package to assess model performance and compute the AUC. Only proteins with data available for more than 95% of participants were included, and missing values were imputed using k-nearest neighbors (KNN) imputation via the VIM package.

We next assessed the predictive capability of MS proteins in estimating baseline tau-PET (neocortical SUVR) and Aβ-PET (Centiloid value). The same training and testing configurations were applied, except that a random forest regressor was used to predict the continuous SUVR and Centiloid value. Model performance was assessed using the Pearson correlation between observed and predicted values. We compared the predictive performance of selected CSF protein panels with conventional biomarkers, including age alone, APOE4 genotype with E4 allele dosage coded as 0, 1, and 2, and CSF Aβ42 and pTau181 as two features. Statistical significance was determined using 100 permutations. The permutations on protein panels and conventional biomarkers were performed in sync, making sure the same permuted training and test participants in each iteration.

## List of Supplementary Materials

**table S1.** Traits of 1,104 participants from the ADNI cohort.

**table S2.** Demographic and clinical characteristics of study participants from the ADNI cohort.

**table S3.** Differential abundance analysis of CSF proteins associated with diagnostic groups (TMT-MS).

**table S4.** Differential abundance analysis of CSF proteins shared between the targeted Selected Reaction Monitoring (SRM) panel and the TMT-MS proteomics cohort across diagnostic groups.

**table S5.** Differential abundance analysis of CSF proteins associated with diagnostic groups (SomaLogic).

**table S6.** Classification performance across 100 iterations of random subsampling to distinguish AT- controls from participants with AD pathology [MCI (due-to-AD) and AD Dementia] using TMT-MS proteomics.

**table S7.** Differential abundance analysis of proteins (using TMT-MS) previously identified as altered along the disease timeline in the DIAN-TU autosomal dominant Alzheimer’s disease cohort.

**table S8.** Classification performance across 100 iterations of random subsampling to distinguish AT- controls from asymptomatic AD using TMT-MS proteomics.

**table S9.** Differential abundance analysis of CSF proteins across Alzheimer’s disease stages (TMT-MS).

**table S10.** Protein panels selected by machine learning models to distinguish AD stages using least absolute shrinkage and selection operator (LASSO).

**table S11.** Differential abundance analysis of TMT-MS proteins correlated with baseline clinical measures of cognition and dementia severity.

**table S12.** Differential abundance analysis of TMT-MS proteins correlated with baseline neuroimaging biomarkers of neurodegeneration and pathology.

## Data Availability

Raw mass spectrometry data and pre- and post-processed plasma protein abundance data and case traits related to this manuscript are available at https://www.synapse.org/Synapse:syn59804727 on the AMP-AD Knowledge Portal, which is a platform for accessing data, analyses and tools generated by the AMP-AD Target Discovery Program and other programs supported by the National Institute on Aging to enable open-science practices and accelerate translational learning. The data, analyses and tools are shared early in the research cycle without a publication embargo on secondary use. ADNI data are available on https://adni.loni.usc.edu/.

## Acknowledgments

This study was supported by the following National Institutes of Health (NIH) funding mechanisms: U01AG061357 (A.I.L. and N.T.S.), RF1AG062181 (N.T.S.), P30AG066511 (A.I.L.), RF1NS139948 (N.T.S.), and R01AG075820 (N.T.S.); as well as the Foundation for the National Institutes of Health (FNIH) AMP-AD 2.0 grant and a grant from the Alzheimer’s Association (ABA-22-974673). Data collection and sharing for this project was funded by the Alzheimer’s Disease Neuroimaging Initiative (ADNI) (National Institutes of Health Grant U01 AG024904) and DOD ADNI (Department of Defense award number W81XWH-12-2-0012). ADNI is funded by the National Institute on Aging, the National Institute of Biomedical Imaging and Bioengineering, and through generous contributions from the following: AbbVie, Alzheimer’s Association; Alzheimer’s Drug Discovery Foundation; Araclon Biotech; BioClinica, Inc.; Biogen; Bristol-Myers Squibb Company; CereSpir, Inc.; Cogstate; Eisai Inc.; Elan Pharmaceuticals, Inc.; Eli Lilly and Company; EuroImmun; F. Hoffmann-La Roche Ltd and its affiliated company Genentech, Inc.; Fujirebio; GE Healthcare; IXICO Ltd.; Janssen Alzheimer Immunotherapy Research & Development, LLC.; Johnson & Johnson Pharmaceutical Research & Development LLC.; Lumosity; Lundbeck; Merck & Co., Inc.; Meso Scale Diagnostics, LLC.; NeuroRx Research; Neurotrack Technologies; Novartis Pharmaceuticals Corporation; Pfizer Inc.; Piramal Imaging; Servier; Takeda Pharmaceutical Company; and Transition Therapeutics. The Canadian Institutes of Health Research is providing funds to support ADNI clinical sites in Canada. Private sector contributions are facilitated by the Foundation for the National Institutes of Health (www.fnih.org). The grantee organization is the Northern California Institute for Research and Education, and the study is coordinated by the Alzheimer’s Therapeutic Research Institute at the University of Southern California. ADNI data are disseminated by the Laboratory for Neuro Imaging at the University of Southern California.

## Funding

This study was supported by the following funding mechanisms:

National Institutes of Health U01AG061357 (AIL and NTS)

National Institutes of Health RF1AG062181 (NTS)

National Institutes of Health P30AG066511 (AIL)

National Institutes of Health RF1NS139948 (NTS)

National Institutes of Health R01AG075820 (NTS)

Foundation for the National Institutes of Health AMP-AD 2.0 (NTS, AIL)

Alzheimer’s Association ABA-22-974673 (NTS, AIL)

## Author contributions

Conceptualization: SR, AIL

Methodology: SR, EBD, NTS, AIL

Investigation: SR, EBD, AS, NTS, AIL

Visualization: SR, NTS, AIL

Funding acquisition: NTS, AIL

Project administration: SR, AIL

Supervision: NTS, AIL

Writing – original draft: SR, NTS, AIL

Writing – review & editing: SR, EBD, AS, FW, DMD, EJF, ECBJ, JJL, NTS, AIL

---

**fig. S1.**
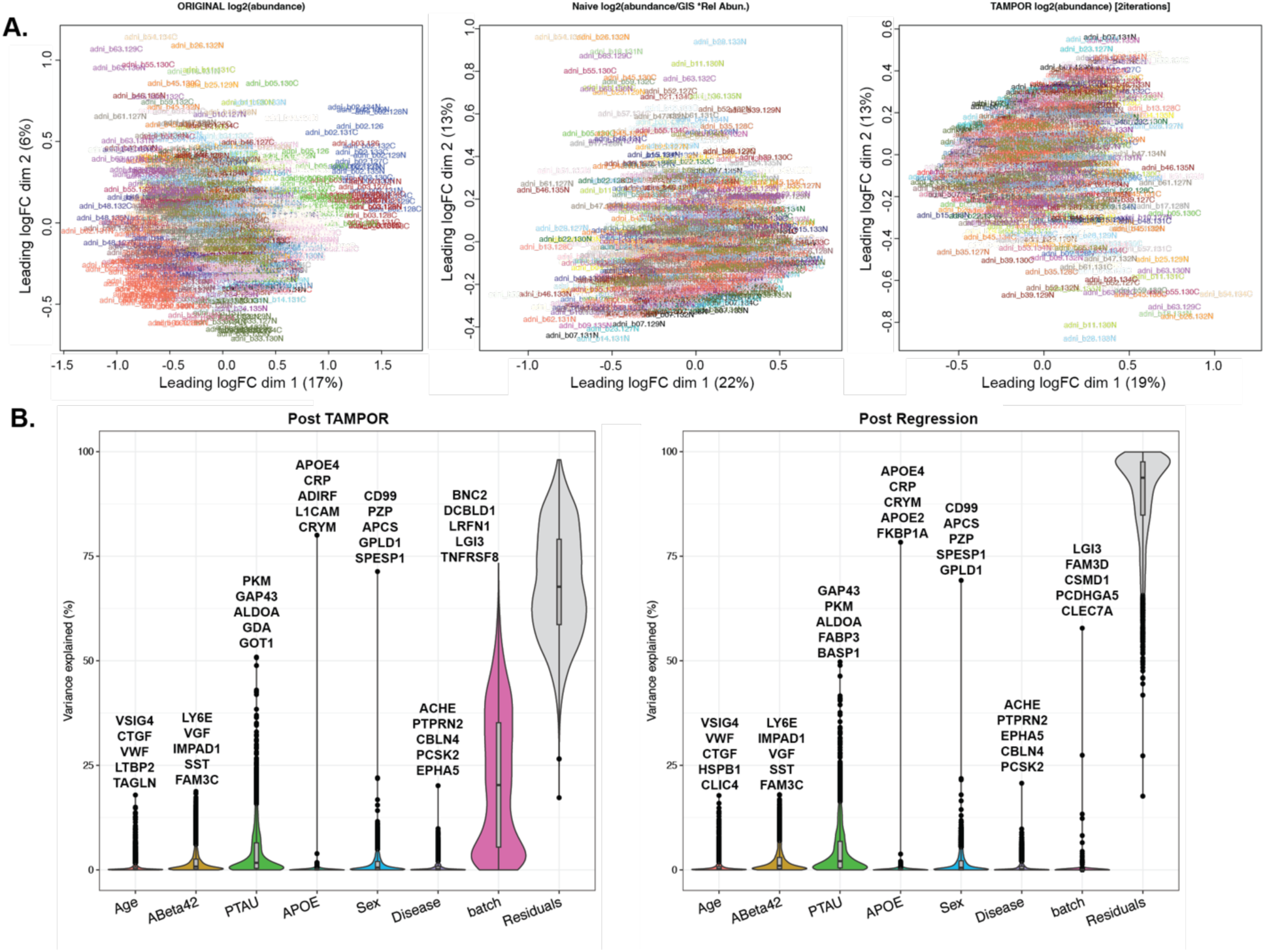
Quality control of the ADNI CSF proteome. (A) Multidimensional scaling (MDS) illustrating TMT-MS batch correction. Log2 abundance, log2 abundance divided by the global internal standard (GIS), and TAMPOR are shown. (B) Variance partition plots were used to visualize the percent variance of each protein in the dataset co-varying with batch, age and sex. The matrix was subjected to bootstrap regression (right) to remove variance due to batch.

**fig. S2.**
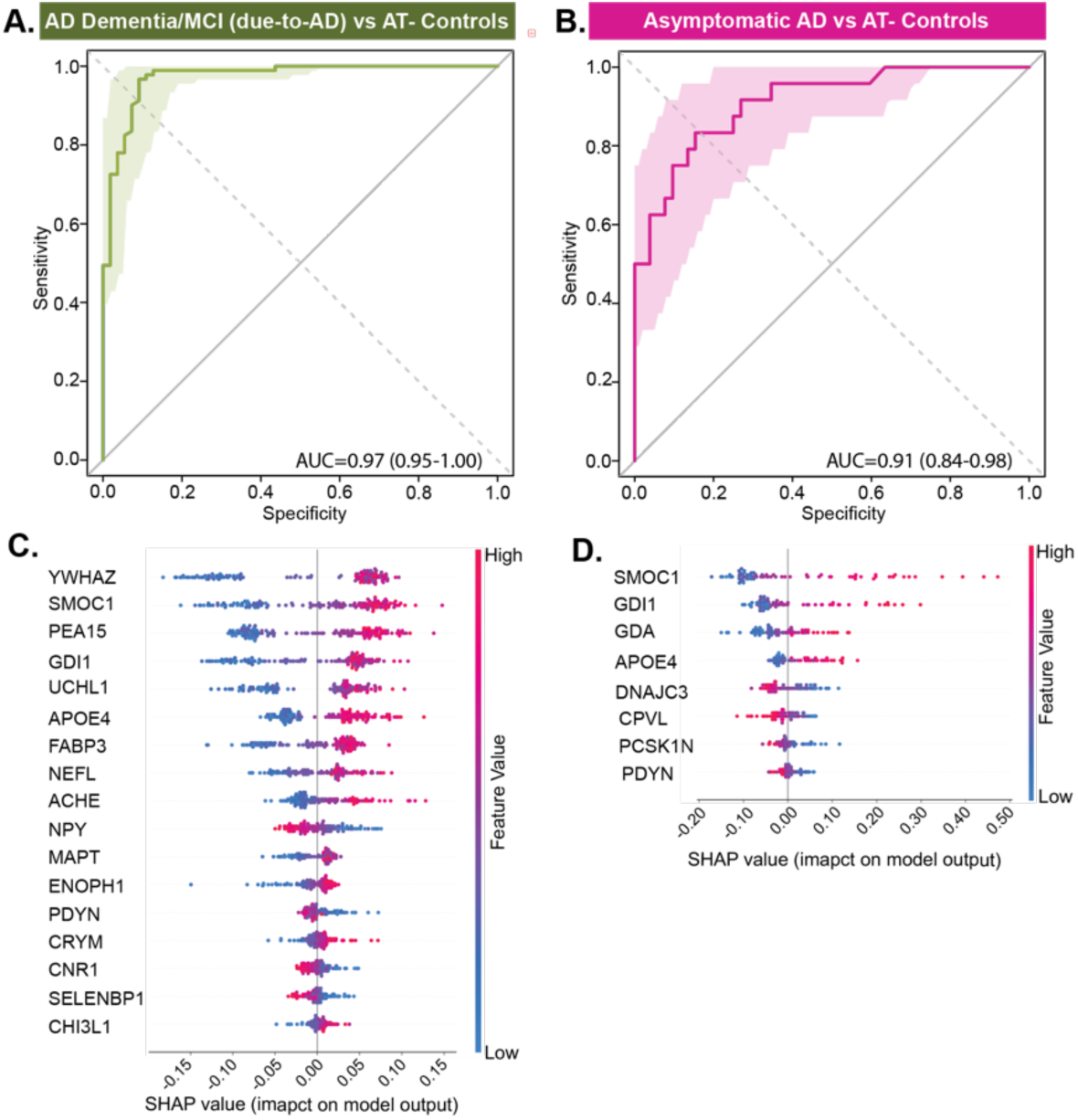
MS proteomics predicts diagnostic groups in ADNI. **(A-B)** MS proteomics data were used to train a classification model. ROC curves from 100 permuted runs illustrate the median Area Under the Curve (AUC) based on an 80–20% train-test split for the following comparisons: (A) AD Dementia/MCI (due-to-AD) versus AT- Controls (AUC=0.97), and (B) asymptomatic AD versus AT- Controls (AUC=0.91). Higher AUC values indicate better classification performance, with values closer to 1.0 reflecting greater sensitivity and specificity. **(C-D)** Top SHAP feature contributions corresponding to the median AUC models shown in A and B are visualized for: (C) AD Dementia/MCI (due-to-AD) versus AT- Controls, and (D) asymptomatic AD versus AT- Controls.

**fig. S3.**
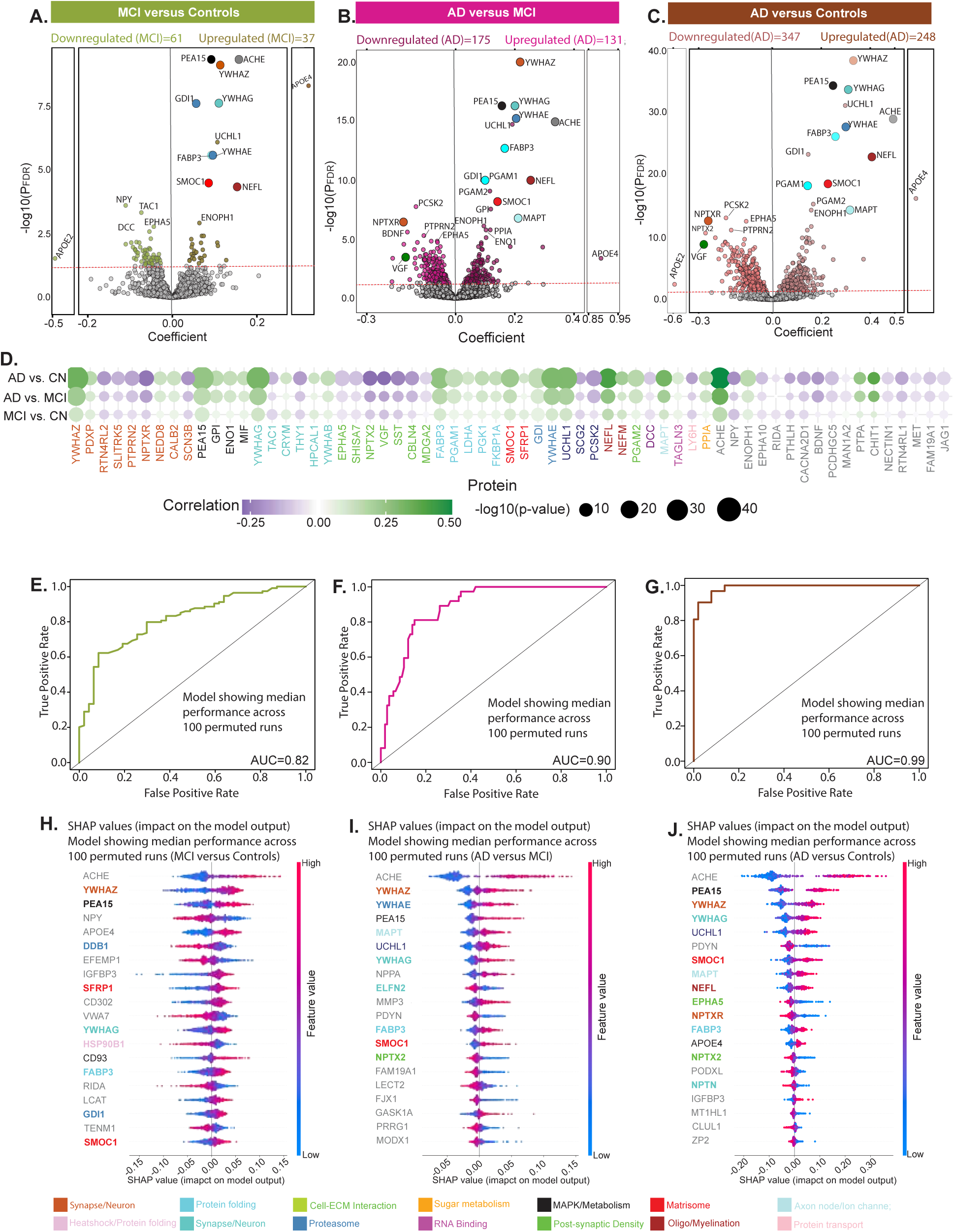
MS proteomics estimates baseline clinical diagnosis in ADNI. **(A-C)** Differential protein expression across diagnostic groups is shown. Volcano plots depict differentially abundant proteins (DAPs) when comparing individuals across diagnostic categories (Controls = 244, MCI = 563, AD = 164): (A) MCI versus Controls, (B) AD versus MCI, and (C) AD versus Controls. Proteins with P_FDR_<0.05 (marked by red line) were considered significant. For clarity, top selected proteins are labeled. **(D)** A heatmap of Pearson correlations is shown for MS proteomics and clinical diagnosis. The MS proteins are labeled as their respective gene symbols, and the strength and direction of correlation is shown by the blue to green color scale. The top 30 proteins with the strongest correlations were selected from each category, and their union is displayed in the heatmaps. **(E-G)** MS proteomics data were used to train a classification model. ROC curves from 100 permuted runs illustrate the median Area Under the Curve (AUC) based on an 80–20% train-test split for the following comparisons: (E) MCI vs. Controls (AUC = 0.82), (F) AD vs. MCI (AUC = 0.90), and (G) AD vs. Controls (AUC = 0.99). Higher AUC values indicate better classification performance, with values closer to 1.0 reflecting greater sensitivity and specificity. **(H-J)** Top SHAP feature contributions corresponding to the median AUC models shown in panels (E-G) are visualized for: (H) MCI vs. Controls, (I) AD vs. MCI, and (J) AD vs. Controls. Colors are assigned to individual proteins names based on their assignment in the brain-derived modules (12).

**fig. S4.**
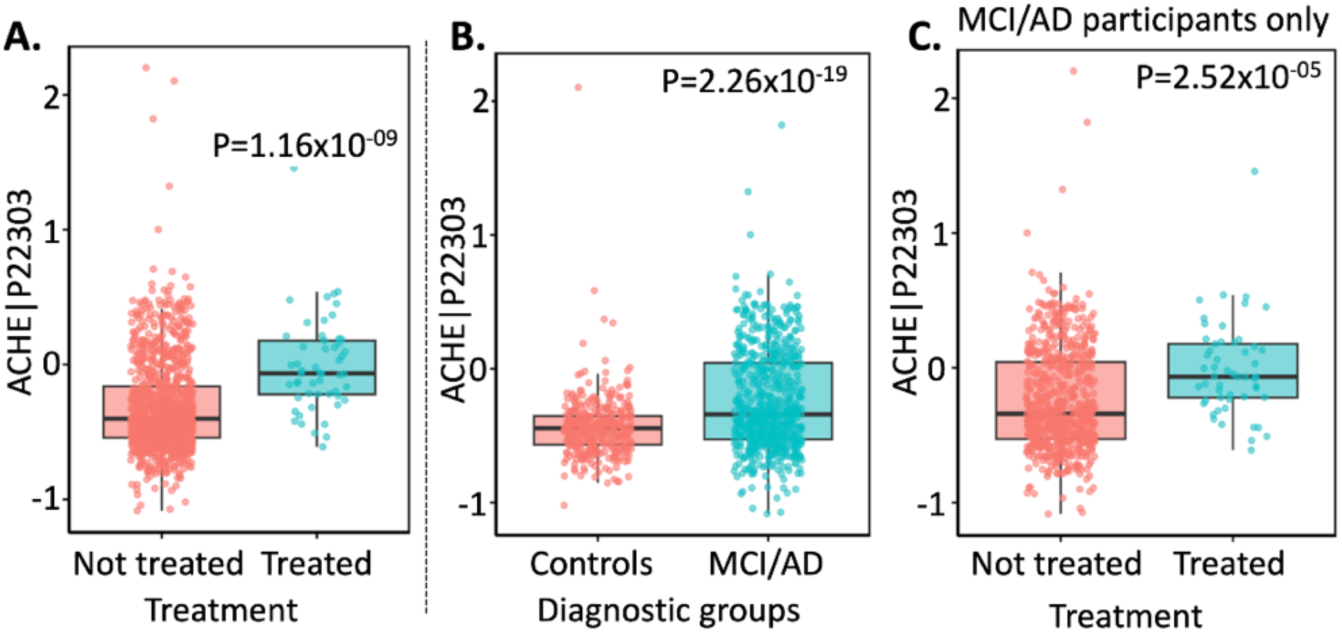
Comparison of AChE expression levels in ADNI participants based on treatment status. Participants treated with Donepezil, Galantamine, Rivastigmine, or Memantine (n=53) and non-treated participants were included in this analysis**. (A)** AChE levels compared between participants treated with cholinergic medications compared to untreated participants prior to CSF collection (P=1.16×10^-9^). **(B)** AChE levels compared between controls and MCI/AD participants regardless of the treatment status (P=2.2×10^-19^). **(C)** AChE levels between treated versus non-treated AD and MCI participants (P=2.52×10^-5^).

**fig. S5.**
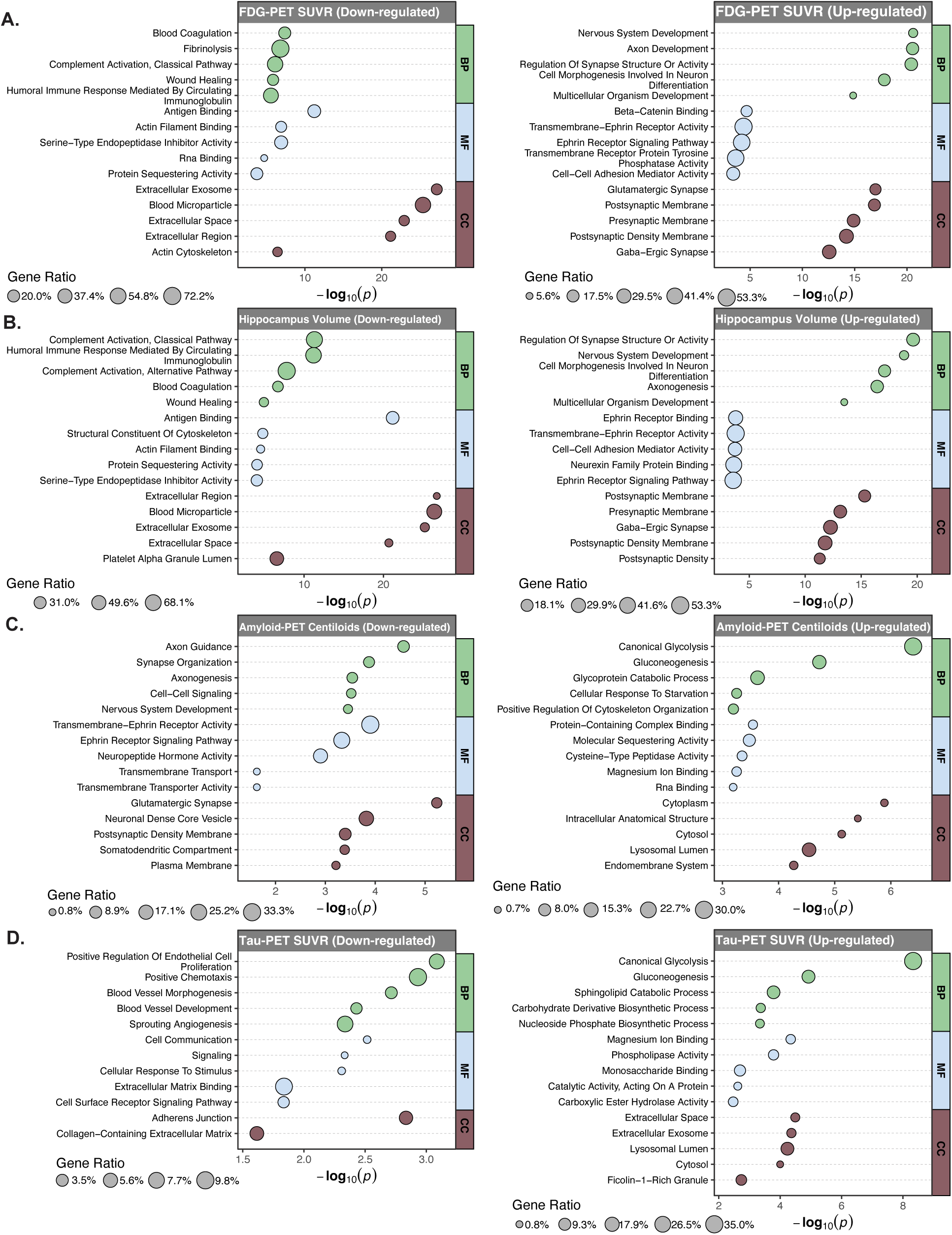
Summary of functional enrichment analyses using Gene Ontology (GO) databases for different categories of proteins. Biological pathways that were up- or downregulated in association with neuroimaging-based biomarkers of **(A-B)** neurodegeneration (hippocampal volume and FDG-PET SUVR) and **(C-D)** pathological burden (tau-PET neocortical SUVR and Aβ-PET Centiloid value) are shown.

